# Accumulation of dihydrosphingolipids and neutral lipids is related to steatosis and fibrosis damage in human and animal models of non-alcoholic fatty liver disease

**DOI:** 10.1101/2022.03.10.22271048

**Authors:** Bohdan Babiy, Bruno Ramos-Molina, Luis Ocaña, Silvia Sacristán, Diego Burgos-Santamaría, Javier Martínez-Botas, Gemma Villa-Turégano, Rebeca Busto, Cristian Perna, M. Dolores Frutos, Agustín Albillos, Óscar Pastor

## Abstract

**Background:** Dihydrosphingolipids are lipid molecules biosynthetically related to ceramides. An increase in ceramides is associated with enhanced fat storage in the liver and inhibition of their synthesis is reported to prevent the appearance of steatosis in animal models. However, the precise association of dihydrosphingolipids with non-alcoholic fatty liver disease (NAFLD) is yet to be established. We employed a diet-induced NAFLD mouse model to study the association between this class of compounds and disease progression.

**Methods:** Mice were fed a high-fat diet enriched in cholesterol and supplemented with glucose and fructose up to 40 weeks. A mouse subgroup was treated with carbon tetrachloride to accelerate fibrosis development. Animals were sacrificed at different time-points to reproduce the full spectrum of histological damage found in human disease, including steatosis (NAFL) and steatohepatitis (NASH) with and without significant fibrosis. Blood and liver tissue samples were obtained from patients (n=195) whose NAFLD severity was assessed histologically. Lipidomic analysis was performed using liquid chromatography-tandem mass spectrometry.

**Results:** Triglyceride, cholesterol ester and dihydrosphingolipid levels were increased in the liver of model mice in association with the degree of steatosis. Dihydroceramide concentrations increased with the histological severity of the disease in liver samples of mice (0.024 ± 0.003 vs 0.049 ± 0.005, non-NAFLD vs NASH-fibrosis, p<0.0001) and patients (0.105 ± 0.011 vs 0.165 ± 0.021, p=0.0221). Several dihydroceramide and dihydrosphingomyelin species were increased in plasma of NAFLD patients and correlated with accumulation of liver triglycerides.

**Conclusions:** Dihydrosphingolipids accumulate in the liver in response to increased free fatty acid overload and are correlated with progressive histological damage in NAFLD. The increase in dihydrosphingolipids is related to upregulation of hepatic expression of enzymes involved in *de novo* synthesis of ceramides.

**HIGHLIGHTS:**

- Neutral lipids and dihydrosphingolipids accumulate in liver in correlation with the histological severity of NAFLD in both mice and humans.
- The ceramide pathway is stimulated to alleviate the free fatty acid excess in liver of NAFLD models.
- Appearance of significant fibrosis is associated with reduced concentrations of neutral lipids but not dihydrosphingolipids in a mouse model of NAFLD.

## INTRODUCTION

Non-alcoholic fatty liver disease NAFLD (also recently designated ’metabolic associated fatty liver disease’, MAFLD) (1) is a growing health issue related to the epidemic of obesity and diabetes in developed countries. The incidence of cirrhosis associated with NAFLD has increased significantly over the years and is considered one of the main causes of chronic liver disease worldwide (2). However, the key drivers leading to predisposition to the development of steatohepatitis (NASH) and fibrosis, the main factors associated with poor outcomes, are currently unknown (3, 4). The lipotoxicity hypothesis proposes that in the early phases of NAFLD (simple steatosis or NAFL), neutral lipids, such as triglycerides (TG) and cholesterol esters (CE), accumulate in the liver in response to the influx of free fatty acids (FFA) from adipose tissue, which corresponds with the appearance of peripheral and hepatic insulin resistance (5). Moreover, FFAs trigger the synthesis of the less abundant but potentially harmful ceramides (Cer), dihydroceramides (dhCer), and other biosynthetically related sphingolipids and dihydrosphingolipids **(Suppl Fig. 1)**. The majority of evidence on the involvement of sphingolipids and dihydrosphingolipids (collectively termed (dihydro)sphingolipids) in NAFLD has been obtained from knockout studies on animals (reviewed in (6, 7)). Earlier studies clearly demonstrate that increase in liver Cer is associated with enhanced FFA uptake, TG storage and impaired glucose utilization in the liver, none of which could be replicated with dhCer (8). Hence, strategies to prevent the increase in liver Cer using inhibitors of serine palmitoyltransferase (*Sptlc*) such as myriocin (9) or blocking the conversion of dhCer to Cer with dihydroceramide desaturase inhibitor (*Degs1*) have been effective in avoiding early steatosis in animal models (10, 11).

Limited studies to date have examined the relationship between (dihydro)sphingolipid accumulation in diet-induced mouse models of NAFLD. Kim et al. (10) reported an increase in the concentration of Cer 34:1 (C16-Cer) in response to a high-fat diet (HFD) enriched in palmitic acid (FA 16:0) at 24 weeks, coincident with increased expression of ceramide synthase (*Cers6*) and associated with higher lipogenesis rates mediated by the nuclear factor SREBP1c (10). In keeping with this finding, the increase in C16-Cer promoted steatosis in the liver, whereas ceramides containing very-long-chain fatty acids (VLCFA) synthetized by *Cers2* isoenzymes (such as Cer 40:1 and Cer 42:1) protected against early NAFL (12). Short-term administration of carbon tetrachloride (CCl_4_), a toxic agent inducing fibrosis progression in mice fed a normal diet, is reported to enhance liver Cer (13). However, studies focusing on the association of (dihydro)sphingolipids with disease progression to NASH and significant stages of fibrosis (NASH-fibrosis) in animal models are lacking.

Similarly, incomplete results on the relationship between (dihydro)sphingolipids and NAFLD progression have been obtained in human studies. A number of earlier investigations reported increased Cer levels in plasma and liver NAFL and NASH patients (14, 15), while recently publications specifically document an increase in dihydrosphingolipids, *−* dihydroceramide (dhCer) and dihydrosphingomyelin (dhSM) *−* in plasma (16, 17) and liver (18, 19). The reliability of conclusions from studies on the involvement of (dihydro)sphingolipids in NAFLD is hampered by the lack of histological representation of the full spectrum of the disease and limited coverage of the dihydrosphingolipid species.

Cross-examination of the roles of sphingolipids and dihydrosphingolipids in relation to the histological course of disease is necessary to understand the involvement of this class of lipids in pathogenesis, which may be of relevance in establishing optimal candidate molecular targets of the ceramide pathway to inhibit progression. The main aim of the current study was to determine the potential associations of (dihydro)sphingolipid changes with histological grading and staging in NAFLD. To address this issue, we compared findings from a diet-induced mouse model of NAFLD and human patients. Our collective results showed that dihydrosphingolipids are more closely related with histological damage than sphingolipids, especially steatosis and therefore neutral lipid accumulation, in both mice and humans.

## METHODS

A full description of the methods is provided in the **Supplementary Methods section**.

### Animal housing conditions

Mice were housed in the animal facility of Hospital Universitario Ramón y Cajal (Num. Reg: ES 280790000092) in a temperature-controlled room under a 12 light/dark cycle with *ad libitum* access to food and water. All interventions were performed during the light cycle in accordance with legislation in Spain (RD 53/2013) and the European Directive (2010/63/EU) and approved by the Ethics Committee of the Ramón y Cajal Hospital, Madrid (ES-280790002001).

### Diets and treatments

Six week-old C57BL/6J male mice received either standard rodent chow or a high-fat diet (HFD) enriched with cholesterol (1.25%) and a high sugar solution of fructose and glucose in drinking water up to 40 weeks. At 22 weeks, a subset of animals received a weekly intraperitoneal injection of CCl_4_ at a final dose of 0.2 μL CCl_4_)/gr of body weight for 6 and 10 weeks. Animals were sacrificed at predetermined time-points (**Suppl. Fig. 2)**. Food was removed 6 h before death. Mice were anesthetized with sevoflorane (2% in oxygen flux at 1 L/min). Blood was drawn by cardiac puncture, the liver extracted, and plasma obtained after centrifugation (1900 g) for biochemistry assays. The median hepatic lobe was preserved in 10% buffered formalin for histological and immunohistochemical analyses.

### Patients

We prospectively enrolled consecutive patients with obesity undergoing bariatric surgery at the two hospital centers (Hospital Clínico Universitario Virgen de la Victoria de Málaga and Hospital Universitario Virgen de la Arrixaca de Murcia, Spain). Inclusion criteria were age range of 18–65 years and obesity of more than 5 years of duration with a body mass index (BMI) ≥40 kg/m^2^.

Additional patients in the plasma lipidomic cohort were recruited from the Gastroenterology Department of Hospital Ramón y Cajal for confirmatory NAFLD diagnosis, suspected based on the presence of steatosis upon abdominal ultrasound examination and/or unexplained aspartate aminotransferase (AST), alanine aminotransferase (ALT) or gamma-glutamyltrasferases (GGT) elevation accompanied by type II diabetes mellitus (T2DM), obesity (BMI) ≥35 kg/m^2^) and metabolic syndrome.

Exclusion criteria were set based on evidence of other causes of liver disease, including viral hepatitis, medication-related disorders, autoimmune disease, hepatocellular carcinoma, haemocromatosis, Wilson’s disease, familial/genetic causes or a previous history of excessive alcohol use (>30 g daily for men and >20 g daily for women) or treatment with any drugs potentially causing steatosis, such as tamoxifen, amiodarone, and valproic acid (20). The study was performed in agreement with the Declaration of Helsinki according to local and national laws and approved by the Ethics and Clinical Research Committees of every center: “Predictive factors of histological lesions in patients with non-alcoholic fatty liver disease” (Hospital Universitario Ramón y Cajal, ref: EC 276-14), “Gut microbiota and related metabolites as potential biomarkers of resolution of non-alcoholic steatohepatitis and improvement of liver fibrosis after bariatric surgery” (Hospital Clínico Universitario Virgen de la Araixaca, ref: 2020-2-4-HCUVA) and “Metabolomic study in obese NAFLD/NASH patients subjected to bariatric surgery” (Hospital Clínico Universitario Virgen de la Victoria de Málaga, ref: S2200002).

### Histology of patient liver biopsies

Intraoperative wedge liver biopsies from bariatric surgery patients at least 1 cm in depth were obtained. One section was snap-frozen and stored at -80°C for lipidomic analysis and the other was formalin-fixed and paraffin-embedded for histological assessment. Percutaneous or transjugular liver biopsy specimens (1 mm in diameter and 1.5 mm in length) were obtained from patients subjected to non-bariatric surgery included in the plasma lipidomic cohort. The minimal set of staining procedures included hematoxylin and eosin (H&E), Masson trichrome, Periodic acid– Schiff (PAS), Perls and reticulin staining. All biopsies were reviewed and scored by trained liver pathologists from the recruitment centers (**Supplementary methods**).

### Histology of mouse liver biopsies

Mouse liver tissue sections were stained with H&E and picrosirius red and evaluated in a blinded manner by an experienced pathologist (P.C). Histological scoring in mice was performed using the SAF (<u>S</u>teatosis, <u>A</u>ctivity and <u>F</u>ibrosis) classification system proposed by Bedossa (21) (Supplementary methods).

### Western blot

Homogenized liver tissue extracts were diluted to 1 mg/mL in lysis buffer (described in **Supplementary methods)** and protein concentrations in the extracts assessed using the bicinchoninic acid assay. For measurement of expression, 40 μg protein was loaded onto 10% SDS gels for electrophoresis and transferred to nitrocellulose membranes. After blocking (0.1% solution of casein in TBS w/v), blots were incubated with primary antibodies for actin alpha 2/α-smooth muscle actin (ACTA2/*α*-SMA; dil 1:1000 v/v), perilipin 2 (PLIN2; dil 1:4000 v/v), sequestosome 1 (p62; dil 1:4000 v/v), collagen type I Alpha 1 chain (Col1A1; dil 1:2000 v/v) and heat shock protein 90 (HSP90; dil 1:2000 v/v), followed by secondary antibodies conjugated to IRDye 800 CW or IRDye 680 LT (LI-COR, Lincoln, NE, USA). Quantification of bands was performed via densitometry using the Odyssey Infrared Imaging System V.3.0 and normalized to the intensity of the loading control (HSP90).

### Real-time PCR gene expression

Homogenized liver tissue was extracted using an RNeasy-Plus Universal Mini Kit and the QIAcube automated purification system (Qiagen, Maryland-USA) according to the manufacturer’s instructions. Following validation, RNA (500 ng) was reverse-transcribed with random hexamers using the PrimeScript RT reagent kit (Takara, California-USA). Real-time PCR amplification was performed on a LightCycler 480 II using the SYBR Green I Master kit (Roche, Basel-Switzerland) on 384-well plates. Data were analyzed using the relative quantification method described by Pfaffl (22), based on the mean values of three housekeeping genes (*18S ribosomal RNA* [*Rn18s], cyclophilin B [Cypb],* and *Human ribosomal protein large P0 [Rplp0]*) as invariant endogenous controls for each sample as described in **Supplementary methods**.

### Oral glucose tolerance test

Oral glucose tolerance test (OGTT) was performed in mice after 5 h of fasting (23) . For this experiment, mice were administered an oral solution of glucose (0.5 g/mL) using a gavage needle (2 g glucose/kg body weight). Blood glucose measurements were conducted on samples (obtained via tail nicking) at 0, 15, 30, 60, and 120 min with the aid of a glucometer (Performa-Accu-Chek. Roche, Basel-Switzerland).

### Blood biochemistry assays

Glucose, AST, ALT, total cholesterol (Chol), total triacylglycerols (Trig), HDL cholesterol (HDLc), insulin, and other blood clinical chemistry measurements were performed using an Architect ci16000 system (Abbott Diagnostics, Illinois-USA). The glycated haemoglobin (HbA1c) level was analyzed using Hb9210 Premier equipment (A. Menarini Diagnostics, Florence-Italy). Free fatty acid (FFA) levels in plasma and liver tissue were measured using a specific ELISA kit (Elabscience, Houston-USA). Procolagen III aminoterminal peptide (PNIIIP), Hyaluronic acid (HA) and metallopeptidase inhibitor 1 (TIMP1) in plasma were analyzed on an ADVIA-Centaur XPT system (Siemens Healthineers, Malvern-USA) and cytokeratin 18 (CK18) evaluated using an ELISA kit (PEVIVA, Stockholm-Sweden).

### Lipidomic analyses of plasma and liver tissue samples

Lipid species were analyzed using targeted liquid chromatography coupled to tandem mass spectrometry following a previously described validated workflow (24). Lipids from homogenized liver tissue (equivalent to 250 µg of total protein) or plasma (50 μL) were extracted following Folch’s methodology (25). A mixture (10 µL) containing one internal standard per lipid class was added to all samples before lipid extraction for molar quantification. Analysis of several batches was necessary for processing on different days. To reduce analytical variability for every batch, we analyzed a self-validated plasma calibrator and three different levels of quality controls. Using this approach, the analytical within-day and between-day relative standard deviations (RSDs) were kept below 20% for the 130 species of the 13 different lipid classes (**Suppl. Fig. 9**).

### Statistical analysis

Statistical analyses were conducted using R software version 3.5.1 (http://www.r-project.org). One-way ANOVA comparisons for quantitative variables between more than two groups were conducted and in cases of significance, post-hoc paired comparisons performed using the Student’s t-test. Comparison between qualitative variables was performed using the Chi-square (χ2) test. P-values of less than 0.05 were considered significant. Correlations between quantitative variables were examined using Pearson’s correlation coefficient. Hierarchical cluster analysis was performed with the aid of k-means algorithm using euclidean distances.

### Data availability

The final datasets used for manuscript preparation are available at Zenodo repository (https://doi.org/10.5281/zenodo.6122303).

## RESULTS

### Metabolic characterization of the NAFLD mouse model

To establish the relationship between (dihydro)sphingolipid species and NAFLD, we initially examined the characteristics of a diet-induced mouse model of NAFLD **(Suppl. Fig. 2**). Mice were fed a HFD enriched in cholesterol, glucose and fructose for up to 40 weeks and ultimately sacrificed at different time points (22, 30 and 40 weeks) to trace disease progression from NAFL to NASH. A subset of animals was treated with CCl_4_, starting at 22 weeks, but kept under the same regime to accelerate progression to higher stages of fibrosis (F3-F4). Animals were sacrificed at 28 weeks (6 injections of CCl_4_) and 32 weeks (10 injections of CCl_4_) for experimental purposes.

HFD feeding increased the weights of animals compared with control littermates (**Fig. 1A**). This increase corresponded to changes in liver weight and liver to body weight ratio (LW/BW) (**Fig. 1B, C**, respectively). Conversely, administration of CCl_4_ induced a decrease in both weight and LW/BW. Transaminase levels in plasma (AST and ALT) were higher from 22 weeks onwards, with a more significant increase at 40 weeks (**Fig. 1D, E**). Administration of CCl_4_ induced a decrease in transaminase levels compared to the HFD-fed group at 22 weeks. Total plasma cholesterol (Chol) and HDL-cholesterol (HDLc) were increased in HFD-fed animals (**Fig. 1F, H**). Surprisingly, plasma triglyceride levels were decreased in the HFD group (**Fig. 1G**). Plasma basal glucose concentrations (Glu) increased from 22 weeks of feeding, showing a decay at 40 weeks or after CCl_4_ administration (**Fig. 1I**). This behavior was compatible with development of insulin resistance (IR) in HFD mice at 22 weeks and confirmed upon glucose challenge (**Fig. 1J, K**). Metabolically, the mouse model showed progressive body weight gain (obesity) accompanied by IR, dyslipidemia (hypercholesterolemia) and elevated levels of biomarkers of hepatocellular damage. All these factors are frequently associated with human NAFLD.

**Fig. 1.**
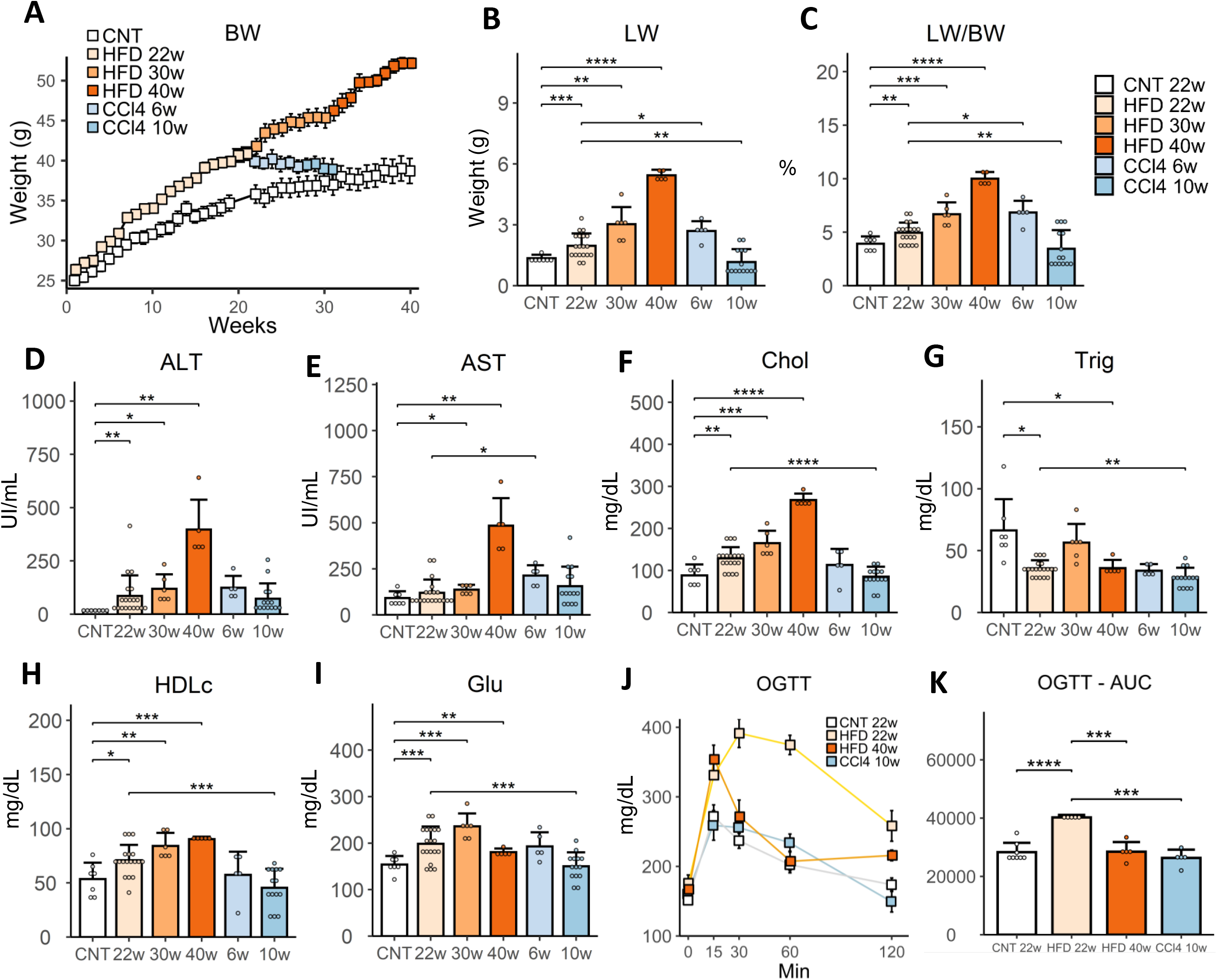
Metabolic characterization of the NAFLD mice model. (A) 8 weeks year old mice were treated with HFD (+ fructose and glucose in water) up to 40 weeks. At 22 weeks a subgroup of mice was treated with CCl_4_ for 6 and 10 weeks. Body weight change during the follow-up period. Normal chow diet for 22 weeks and 30 weeks (CNT, n=14); HFD for 22 weeks (HFD 22w, n=19); HFD for 30 weeks (HFD 30w, n=6); HFD for 40 weeks (HFD 40w, n =5), HFD 22w+CCl_4_ for 6 weeks (CCl_4_ 6w, n=5), HFD 22w+CCl_4_ for 10 weeks (CCl_4_ 10w, n=14). (B) Liver weight (LW). (C) liver per body weight ratio (LW/BW). (D) Plasma ALT. (E) Plasma AST. (F) Plasma total cholesterol. (G) Plasma total triglycerides. (H) Plasma high-density lipoprotein cholesterol. (I) Fasting blood glucose. (J) Oral glucose tolerance test. (H) Area under the curve in the OGTT. Results were expressed as mean ± SD. For better clarity, only the student t-test comparison respect to the CNT group for HFD 22w, HFD 30w, and HFD 40w and CCl_4_ 6w, CCl_4_ 10w respect to HFD 22w, are showed. *p <0.05, **p <0.01, ***p <0.001, **** p<0.0001. ALT, alanine aminotransferase; AST, aspartate aminotransferase; CCl_4_, carbon tetrachloride; HFD, high fat diet; CNT, normal chow diet; Chol, total cholesterol; Trig, total triglycerides; Glu, glucose; HDLc, high-density lipoprotein cholesterol; OGTT, oral glucose tolerance test; AUC, area under the curve, LW, Liver weight, BW, body weight.

### Mouse liver histology

Representative images of livers, H&E - and sirius red-stained sections are presented in **Fig. 2**. At 22 weeks, HFD mice showed clear signs of neutral lipid accumulation surrounding the central vein (zone 3) with predominance of microvesicular steatosis, which progressed to macrosteatosis from 30 weeks onwards. An increase in inflammatory activity was evident at 30 weeks, manifested by the increased presence of clusters of inflammatory cells and hepatic crown-like structures (hCLS) associated with greater staining of aortic smooth muscle actin (ACTA2/*α*-SMA) (**Suppl. Fig. 3**). However, ballooned hepatocytes, a hallmark of NASH in human NAFLD, were only detected at 40 weeks, concordant with the steep elevation of transaminase levels in plasma (**Fig. 1D, E**), and also observed in mice treated with CCl_4_. Mild signs of perisinusoidal stage F1 fibrosis were evident at 30 weeks in 5 out of 7 animals, whereas at 40 weeks, all mice showed progression to perisinusoidal and periportal stage F2 fibrosis. All mice treated with CCl_4_ exhibited stage F *≥* 2, with progression to stage 3 after 10 weeks, accompanied by lower steatosis scores, severe necroinflammatory damage and formation of regenerative micronodules. For each biopsy, a SAF score summarizing the main histological lesions was obtained (**Suppl. Fig 4A**).

**Fig. 2.**
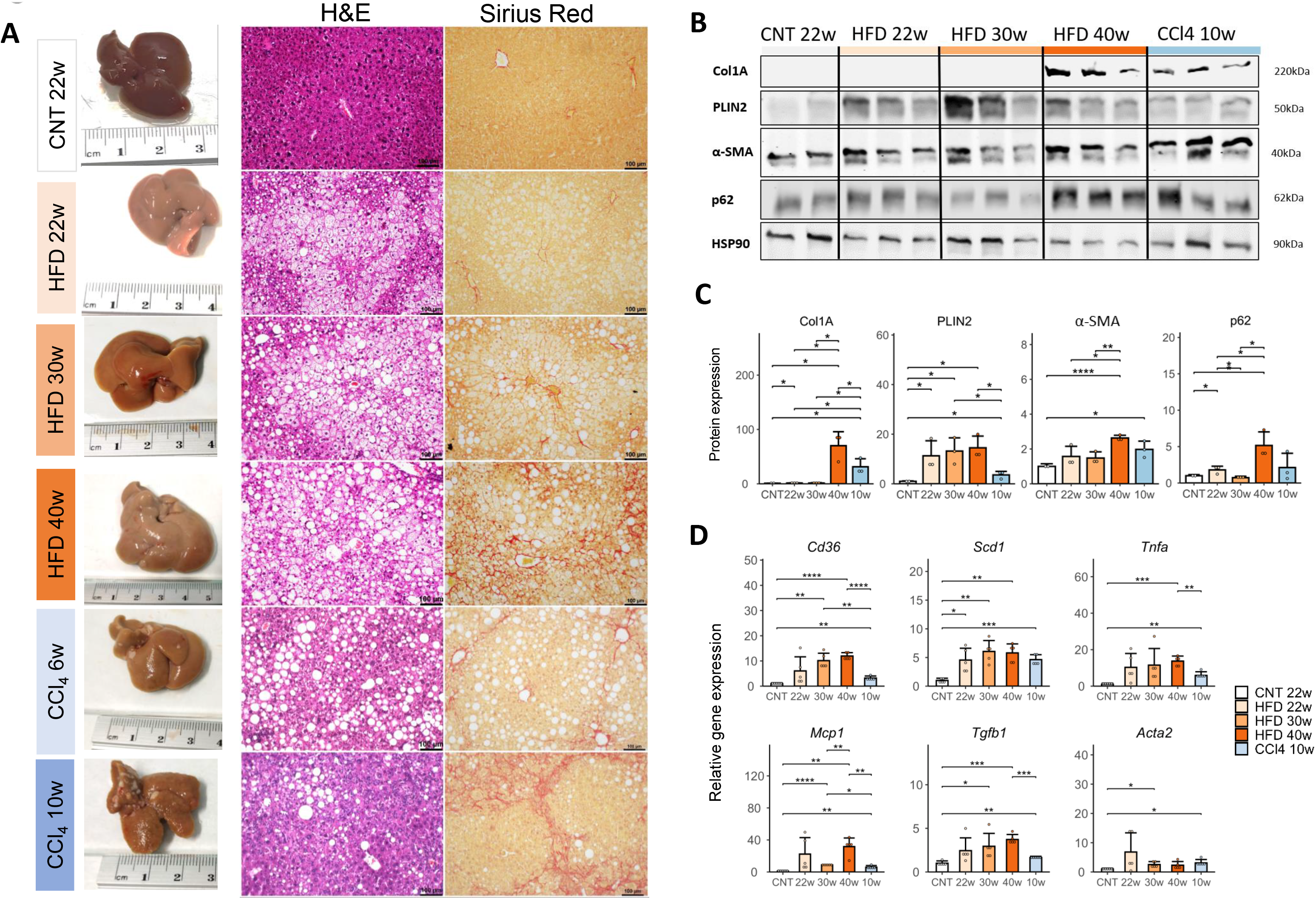
Histological features correlate with protein and gene expression biomarkers or disease progression in the mice model. (A) Macroscopic appearance, hematoxylin and eosin, and sirius red staining of representative liver from mice on normal chow diet for 22 weeks (CNT 22w); high-fat diet HFD for 22 weeks (HFD 22w); HFD for 30 weeks (HFD 30w); HFD for 40 weeks (HFD 40w), HFD 22w+CCl_4_ for 6 weeks (CCl_4_ 6w) and HFD 22w+CCl_4_ for 10 weeks (CCl_4_ 10w). Original magnification x100. (B) Representative Western-blot of PLIN2, *α*-SMA, p62, COLA1A and housekeeping standard HSP90 in liver lysates from CNT 22w, HFD 22w, HFD 30w, HFD 40w, CCl_4_ 10w. (C) Densitometric analysis of the ratio of PLIN2, *α*-SMA, p62 and COLA1A, normalized by HSP90. Data represent the mean ± SD of n=5 samples per condition. (C) Gene expression levels of *Cd36, Scd1, Tnfa* and *Tfgb1* in the liver of CNT 22w, HFD 22w, HFD 30w, HFD 40w, CCl_4_ 10w. Data represent the mean ± SD of n=5 samples per condition. *p <0.05, **p <0.01, ***p <0.001. ****p <0.0001.

Western blot analysis of protein expression of perilipin-2 (PLIN2, a marker of lipid droplet formation), ACTA2/*α*-SMA (a marker of stellate cell activation), p62 (a marker of Mallory-Denk bodies) and collagen alpha-1(I) chain (COL1A1) associated with scar tissue formation confirmed our histological findings (**Fig. 2B, C**). In a similar manner, expression patterns of genes related to fatty acid transport and synthesis (*Cd36, Scd1*), inflammation (*Tnfa, Mcp1*) and fibrogenesis (*Acta2*, *Tgfb1*) corroborated with the changes observed in the model (**Fig. 2D**).

### Time-course analysis of plasma dihydro(sphingolipid) changes in NAFLD mice

Metabolic and histological findings, together with protein and gene expression changes, confirmed the suitability of the mouse model for investigating the relationship between (dihydro)sphingolipids and progression of NAFLD. To determine the associations of these compounds with NAFLD development, changes in the plasma concentrations were monitored over time. Dihydrosphingolipid classes (dhCer and dhSM) were increased early in mouse plasma only after 4 weeks of HFD feeding and remained higher in the HFD-fed group during the whole follow-up period. However, the increase in sphingolipids was less significant and Cer and SM areas under the curves (AUC) were not markedly different from those of control mice (**Fig 3A, B**). The increase in dihydrosphingolipids was coincident with that of FFA in plasma and liver tissue (**Fig. 3C**).

**Fig. 3.**
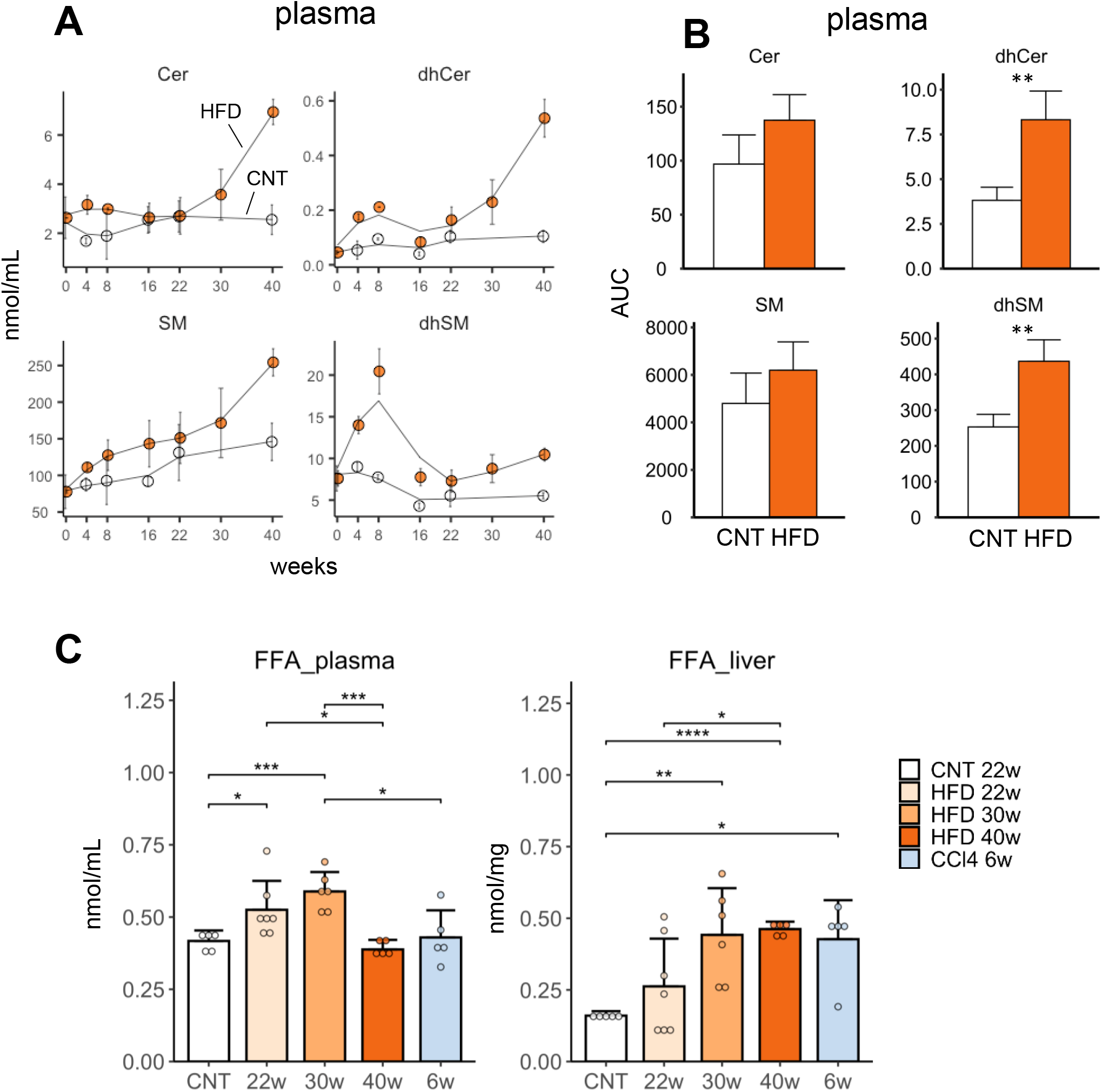
Dihydrosphingolipid evolution is associated to the increase in FFA in the mice model. (A) Time-course of plasma sphingolipid classes: Cer, dhCer, SM, dhSM in normal chow (CNT) and high-fat diet (+fructose and glucose). Lipid classes were calculated as the sum composition from the corresponding lipid species. Data represent the mean ± SEM of n=3-4 animals per group. (B) AUC of sphingolipid molecular species. Data represent the mean ± SEM of n=3-4 animals per group. (C) FFA in plasma and liver tissue samples from mice on normal chow diet for 22 weeks (CNT 22w); high-fat diet HFD for 22 weeks (HFD 22w); HFD for 30 weeks (HFD 30w); HFD for 40 weeks (HFD 40w), HFD 22w+CCl_4_ for 6 weeks (CCl_4_ 6w). Data represent the mean ± SD of n=5-7 animals per group. ** p<0.01. Cer, ceramide; dhCer, dihydroceramide; SM, sphingomyelin; dhSM, dihydrosphingomyelin. FFA, free fatty acids.

### Analysis of liver dihydrosphingolipids of the NAFLD mouse model

While time of diet and exposure to CCl_4_ determined disease progression in our mouse model, we observed substantial heterogeneity in liver histology when these factors were solely considered (**Suppl. Fig. 4A)**. Accordingly, mice were further categorized based on histological findings into non-NAFLD (normal diet-fed), NAFL, NASH (steatohepatitis without significant fibrosis, F0-F1), and NASH-fibrosis (steatohepatitis with significant fibrosis, F2-F4) groups (**Suppl. Fig. 4B**). This grouping system appears the most clinically relevant for NAFLD prognosis (3, 4) and facilitates comparison with human NAFLD histology.

As expected, lipidomic analysis of mouse livers revealed an increase in TG and CE species in NAFLD mice in concordance with the steatosis score (**Fig. 4A**). Additionally, the concentration of 1-o-acylceramide (ACer), a lipid derived from condensation of fatty acids to one of the free hydroxyl group of ceramides, was higher, although very low relative to that of TG (**Suppl. Table S1**). The values obtained for the dihydrosphingolipid classes (dhCer, dhSM and dhHexCer) were higher in NAFLD compared to non-NAFLD mice, especially those corresponding to very-long-chain fatty acid species (FA 22:0, FA 24:0 and FA 24:1) (**Fig. 4B**). We did not observe a clear trend of increase for Cer although the concentration was higher in the NASH group, whereas SM was specifically increased in liver of the NASH-fibrosis group. Advanced fibrosis (F3-F4) was associated with reduced accumulation of TG, CE and ACer but higher levels of dihydrosphingolipids (**Suppl. Fig. 5**).

**Fig. 4.**
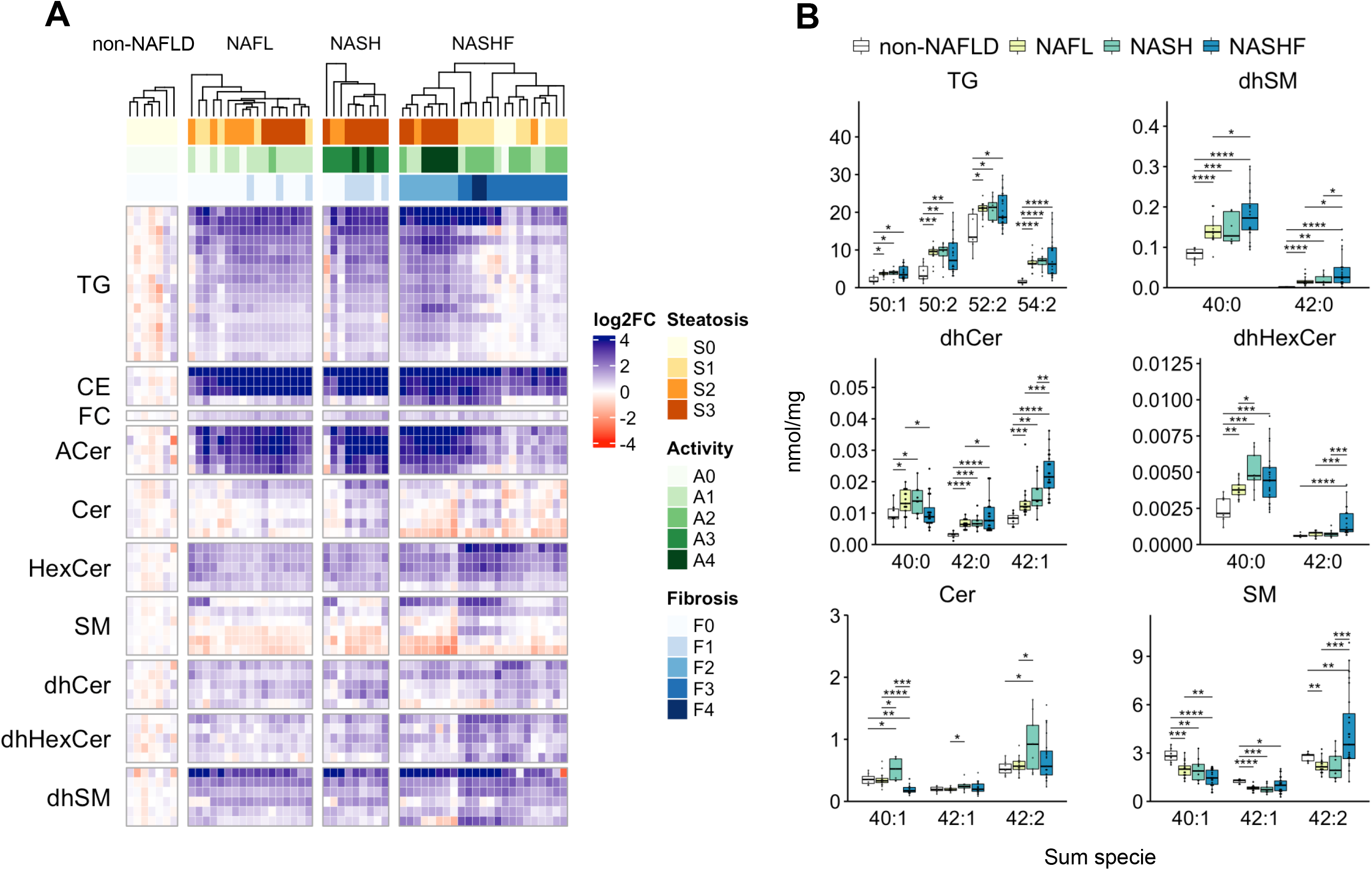
Lipidomic analysis of (dihydro)sphingolipids in NAFLD mice liver. (A) Quantitative analysis of lipid species from 10 classes (see Suppl. Table 2 for class concentrations), were determined in livers of mice histologically classified as: non-NAFLD (n=7), simple steatosis (NAFL, n=17), steatohepatitis without significant fibrosis, stage of fibrosis F0-F1 (NASH, n=9) and steatohepatitis + stage of fibrosis F2-F4 (NASH-fibrosis, n=24). The values of Log_2_-fold change of each animal (column) and each lipid specie (row) is plotted in the in the heatmap and ordered by the hierarchical clustering of individuals (columns). (B) Hepatic concentrations of representative species of TG, dhSM, dhCer, dhHexCer, Cer and SM. Average values are represented as boxplots, with points showing the value of each mice. *p <0.05, *p <0.01, ***p <0.001 and ****p <0.001, using Student’s t-test. TG, triglyceride; CE, Cholesteryl ester; FC, free cholesterol; ACer, 1-o-Acylceramide; Cer, ceramide; HexCer, hexosylceramide; SM, Sphingomyelin; dhCer, dihydroceramide; dhHexCer, dihydrohexosylceramide dhSM, dihydrosphingomyelin. NASF; NASH-fibrosis

We examined the hypothesis that the increase in dihydrosphingolipids in liver may be a result of changes in the expression of enzymes implicated in their synthesis. In keeping with our supposition, hepatic gene expression of enzymes involved in *de novo* sphingolipid synthesis, for instance, serine palmitoyl transferases (*Sptlc1* and *Sptlc2*), ceramide synthases (*Cers2-6*), dihydroceramide desaturase-1 (*Degs1*) and sphingomyelin synthases (*Sgms1* and *Sgms2*), increased with severity of NAFLD (**Suppl. Fig. 6**).

### Lipidomic analysis of liver tissue of NAFLD patients

To further explore whether the changes in (dihydro)sphingolipid levels in mouse liver could be replicated in human NAFLD, we analyzed the concentrations of lipid species in liver from 94 severe or morbidly obese patients subjected to bariatric surgery with NAFLD diagnosis established via histology. The clinical, biochemical and demographic characteristics of the cohort are presented in **Table 1** and the SAF scores in **Suppl. Fig. 7A**. Average age and BMI were determined as 48.1 ± 9.2 years and 46.4 ± 6.7 kg/m^2^, with a female predominance (70%). Liver histology revealed some degree of NAFLD in 73 participants (78%). The rates of hyperlipidemia, hypertension and T2DM were higher in NAFLD than non-NAFLD groups, specifically in cases of NASH-fibrosis (**Table 1**). The results of glycemic parameters (Glu, HbA1c, homeostatic model assessment for insulin resistance [HOMA-IR]) confirmed a worsening in the metabolic condition, particularly in the NASH-fibrosis group. ALT displayed a slight increase under conditions of significant fibrosis, whereas the plasma fibrosis biomarkers, such as hyaluronic acid (HA), cytokeratin-18 (CK18) and tissue inhibitor metallopeptidase-1 (TIMP1), and fibrosis scores (ELF, FIB-4 and NFS) were slightly increased with disease progression and aligned with severity of disease (**Table 1**).

**Table 1.** Clinical and biochemical values in the bariatric surgery cohort. Patients were classified according to their liver histological findings into four groups: non-NAFLD, simple steatosis (NAFL), steatohepatitis without significant fibrosis, stage of fibrosis F0-F1 (NASH) and steatohepatitis + stage of fibrosis F2-F4 (NASH-fibrosis). Values are expressed in % (N/N total in group) or mean ± SD or (N total in group). (a) Significant, p<0.05 from non-NAFLD. (b) Significant, p<0.05 from NAFL. (c) Significant, p<0.05 from NASH. (d) Significant, p<0.05 from NASH-fibrosis.

| Variable | non-NAFLD | NAFL | NASH | NASH-fibrosis |
| --- | --- | --- | --- | --- |
| **Demographics** |  |  |  |  |
| Number, % | 22 (21/94) | 47 (44/94) | 16 (15/94) | 15 (14/94) |
| Female, % | 76 (16/21) | 70 (31/44) | 87 (13/15) | 43 (6/14) |
| Hypertension, % | 29 (6/21) | 55 (24/44) | 40 (6/15) | 71 (10/14) |
| T2DM, % | 29 (6/21) | 52 (23/44) | 47 (7/15) | 71 (10/14) |
| Hyperlipidaemia, % | 33 (7/21) | 66 (29/44) | 60 (9/15) | 71 (10/14) |
| **Anthropometry** |  |  |  |  |
| Age, y | 43.1 ± 2.2 (21)<sup>b,c,d</sup> | 49.2 ± 1.3 (44)<sup>a</sup> | 49.0 ± 2.3 (15)<sup>a</sup> | 51.071 ± 2.6 (14)<sup>a</sup> |
| Weight, Kg | 121.7 ± 4.5 (21)<sup>d</sup> | 126.9 ± 3.5 (44)<sup>d</sup> | 119.3 ± 6.5 (15)<sup>d</sup> | 147.3 ± 8.4 (14)<sup>a,b,c</sup> |
| BMI, Kg/m<sup>2</sup> | 44.5 ± 1.3 (21)<sup>d</sup> | 46.5 ± 1.1 (44) | 45.7 ± 1.7 (15) | 49.4 ± 1.7 (14)<sup>a</sup> |
| **Laboratory tests** |  |  |  |  |
| Glucose, mg/dL | 91.3 ± 4.0 (20)<sup>b,c,d</sup> | 108.3 ± 4.1 (43)<sup>a,d</sup> | 113.7 ± 8.5 (15)<sup>a</sup> | 133.9 ± 9.0 (14)<sup>a,b</sup> |
| HbA1c, | 5.4 ± 0.1 (19)<sup>b,c,d</sup> | 6.2 ± 0.2 (40)<sup>a,d</sup> | 6.4 ± 0.4 (15)<sup>a</sup> | 7.3 ± 0.4 (14)<sup>a,b</sup> |
| HOMA-IR | 2.2 ± 0.3 (20)<sup>b,c,d</sup> | 4.3 ± 0.4 (38)<sup>a,d</sup> | 4.2 ± 0.8 (11)<sup>a,d</sup> | 7.2 ± 1.2 (12)<sup>a,b,c</sup> |
| Insulin, μU/mL | 10.3 ± 1.5 (21) | 16.5 ± 1.5 (38) | 15.4 ± 2.7 (11) | 21.2 ± 3.3 (12) |
| ALT, IU/mL | 30.2 ± 6.7 (20)<sup>d</sup> | 27.5 ± 2.3 (41) | 32.1 ± 4.1 (15) | 44.9 ± 7.6 (14)<sup>a</sup> |
| AST, IU/mL | 24.7 ± 6.0 (19)<sup>d</sup> | 24.4 ± 1.5 (43) | 30.8 ± 4.4 (15) | 33.1 ± 3.6 (14)<sup>a</sup> |
| GGT, IU/mL | 21.2 ± 4.4 (20)<sup>c</sup> | 27.3 ± 3.3 (41) | 46.7 ± 11.3 (15)<sup>a</sup> | 42.6 ± 11.7 (14) |
| Triglycerides, mg/dL | 107.2 ± 9.0 (19)<sup>b,c,d</sup> | 165.7 ± 13.0 (41)<sup>a</sup> | 166.9 ± 18.5 (15)<sup>a</sup> | 186.9 ± 36.1 (14)<sup>a</sup> |
| Cholesterol, mg/dL | 164.4 ± 8.0 (20) | 171.4 ± 6.2 (42) | 168.1 ± 10.5 (15) | 181.8 ± 10.9 (13) |
| HDLc, mg/dL | 45.4 ± 1.9 (20) | 40.8 ± 1.7 (40) | 40.8 ± 2.4 (15) | 41.8 ± 2.6 (14) |
| LDLc, mg/dL | 102.3 ± 6.8 (18) | 99.2 ± 5.7 (41) | 93.6 ± 9.7 (15) | 103.6 ± 8.4 (14) |
| Albumin, g/dL | 3.9 ± 0.1 (21) | 4.0 ± 0.1 (44) | 4.0 ± 0.1 (15) | 3.9 ± 0.1 (14) |
| Platelet, $\times 10^9$/L | 273.8 ± 14.1 (21) | 270.1 ± 12.6 (43) | 274.2 ± 21.7 (14) | 244.4 ± 17.1 (14) |
| **Fibrogenesis tests** |  |  |  |  |
| HA, ng/mL | 25.8 ± 5.5 (13)<sup>d</sup> | 35.1 ± 5.3 (24) | 41.5 ± 11.7 (6) | 63.0 ± 16.6 (14)<sup>a</sup> |
| PIIINP, ng/mL | 4.9 ± 0.4 (13)<sup>b</sup> | 6.0 ± 0.4 (24)<sup>a</sup> | 8.8 ± 3.3 (6) | 6.2 ± 0.7 (14) |
| TIMP, ng/mL | 124.2 ± 9.8 (13)<sup>d</sup> | 137.4 ± 5.5 (24)<sup>d</sup> | 141.5 ± 14.1 (6) | 180.9 ± 17.1 (14)<sup>a,b</sup> |
| CK18, U/L | 135.5 ± 21.8 (14)<sup>d</sup> | 127.9 ± 12.6 (24)<sup>d</sup> | 124.3 ± 17.4 (6)<sup>d</sup> | 299.2 ± 52.3 (14)<sup>a,b,c</sup> |
| ELF score | $7.9 \pm 0.3$ (13)<sup>d</sup> | $8.4 \pm 0.1$ (24) | $8.7 \pm 0.4$ (6) | $8.9 \pm 0.2$ (14)<sup>a</sup> |
| FIB-4 score | $0.691 \pm 0.077$ (19)<sup>b,d</sup> | $0.964 \pm 0.076$ (40)<sup>a</sup> | $1.082 \pm 0.205$ (14) | $1.12 \pm 0.127$ (14)<sup>a</sup> |
| NFS score | $-0.835 \pm 0.317$ (19)<sup>b,d</sup> | $0.015 \pm 0.258$ (40)<sup>a</sup> | $-0.148 \pm 0.453$ (14) | $0.703 \pm 0.325$ (14)<sup>a</sup> |
| ***Liver lipid classes*** |  |  |  |  |
| TG, nmol/mg | $30.1 \pm 2.6$ (21)<sup>b,c,d</sup> | $68.3 \pm 3.8$ (44)<sup>a,c,d</sup> | $80.5 \pm 5.8$ (15)<sup>a,b</sup> | $96.4 \pm 7.6$ (14)<sup>a,b</sup> |
| CE, nmol/mg | $14.3 \pm 1.3$ (21)<sup>b,c,d</sup> | $23.8 \pm 1.6$ (44)<sup>a,c,d</sup> | $35.7 \pm 4.4$ (15)<sup>a,b</sup> | $29.9 \pm 3.2$ (14)<sup>a,b</sup> |
| FC, nmol/mg | $54.7 \pm 1.6$ (21)<sup>b,d</sup> | $59.5 \pm 1.8$ (44)<sup>a,d</sup> | $54.6 \pm 2.4$ (15)<sup>d</sup> | $66.2 \pm 1.9$ (14)<sup>a,b,c</sup> |
| ACer, nmol/mg | $0.006 \pm 0.001$ (21)<sup>b,c,d</sup> | $0.019 \pm 0.001$ (44)<sup>a,c,d</sup> | $0.032 \pm 0.003$ (15)<sup>a,b,d</sup> | $0.047 \pm 0.005$ (14)<sup>a,b,c</sup> |
| Cer, nmol/mg | $1.4 \pm 0.1$ (21) | $1.6 \pm 0.1$ (44) | $1.7 \pm 0.2$ (15) | $1.7 \pm 0.1$ (14) |
| HexCer, nmol/mg | $0.48 \pm 0.03$ (21)<sup>b,c,d</sup> | $0.57 \pm 0.02$ (44)<sup>a</sup> | $0.57 \pm 0.04$ (15)<sup>a</sup> | $0.58 \pm 0.04$ (14)<sup>a</sup> |
| SM, nmol/mg | $17.3 \pm 0.6$ (21) | $17.3 \pm 0.6$ (44) | $17.2 \pm 0.8$ (15) | $17.6 \pm 0.7$ (14) |
| dhCer, nmol/mg | $0.105 \pm 0.011$ (21)<sup>c,d</sup> | $0.113 \pm 0.007$ (44)<sup>d</sup> | $0.147 \pm 0.019$ (15)<sup>a</sup> | $0.165 \pm 0.021$ (14)<sup>a,b</sup> |
| dhHexCer, nmol/mg | $0.007 \pm 0.001$ (21)<sup>d</sup> | $0.007 \pm 0.001$ (44)<sup>d</sup> | $0.008 \pm 0.001$ (15) | $0.010 \pm 0.001$ (14)<sup>a,b</sup> |
| dhSM, nmol/mg | $3.46 \pm 0.32$ (21)<sup>d</sup> | $3.26 \pm 0.19$ (44)<sup>d</sup> | $3.56 \pm 0.36$ (15)<sup>d</sup> | $4.88 \pm 0.49$ (14)<sup>a,b,c</sup> |
| LPC, nmol/mg | $1.12 \pm 0.09$ (21) | $1.12 \pm 0.05$ (44)<sup>d</sup> | $1.11 \pm 0.09$ (15) | $0.95 \pm 0.06$ (14)<sup>a</sup> |
| PC, nmol/mg | $36.9 \pm 2.7$ (21)<sup>c,d</sup> | $35.6 \pm 1.9$ (44)<sup>c,d</sup> | $54.9 \pm 4.4$ (15)<sup>a,b,d</sup> | $29.5 \pm 1.8$ (14)<sup>a,b,c</sup> |
| PE, nmol/mg | $66.6 \pm 4.3$ (21)<sup>b</sup> | $57.6 \pm 2.4$ (44)<sup>a,d</sup> | $58.7 \pm 3.9$ (15) | $66.0 \pm 3.2$ (14)<sup>b</sup> |

T2DM. Type II diabetes mellitus ; HbA1c, Glycated haemoglobin; HOMA-IR, homeostatic model
assessment for insulin resistance; ALT, alanine aminotransferase; AST, aspartate aminotransferase;
GGT, Gamma glutamyltransferase; BMI, body mass index; HDLc, high-density lipoprotein
cholesterol; LDLc, Low-density lipoprotein cholesterol; HA, Hyaluronic Acid; PIIINP, Procollagen
III aminoterminal peptide; TIMP1, Tissue inhibitor of metalloproteinase-1; CK18, Apoptosis-
associated caspase-cleaved keratin-18. ELF, Enhanced liver fibrosis score; FIB-4, Fibrosis-4 index;
NFS, NAFLD fibrosis score.

Importantly, although non-NAFLD control livers were obtained from obese individuals without histologically proven disease and not truly healthy subjects, the lipidomic analysis of liver revealed a significant increase in neutral lipids (TG, CE and ACer) in NAFLD relative to non-NAFLD groups (**Fig. 5A** & **Table 1**). Moreover, dhCer, dhSM and dhHexCer, and in particular, lipid molecules containing very-long-chain fatty acids (**Fig. 5B**), showed a gradual increase from NAFL to NASH-fibrosis conditions. These results in human liver were similar to that observed with the mouse model. Levels of Cer and dhCer species correlated well with the values of TG species (**Suppl. Fig. 8)**. Likewise, we observed no differences in Cer and SM contents among NAFL, NASH and NASH-fibrosis patient groups (**Table 1**). A significant correlation was evident in the dihydrosphingolipid contents, especially dhCer, but not other sphingolipid classes (exemplified by Cer) between liver and plasma (**Fig. 5C**).

**Fig. 5.**
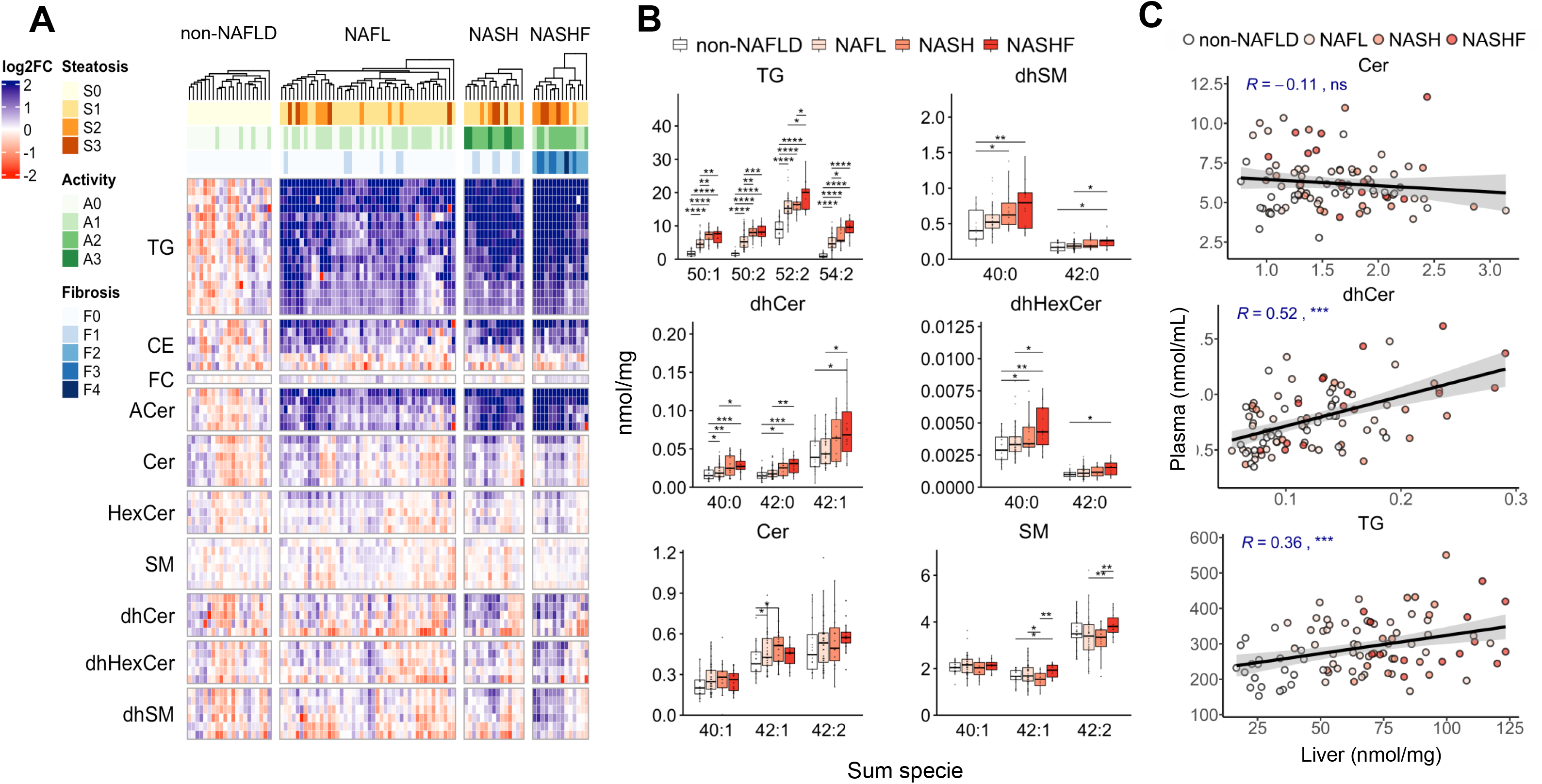
Lipidomic analysis of (dihydro)sphingolipids of livers from the bariatric surgery cohort. (A) Quantitative analysis of lipid species from 10 classes (see Table 1 for lipid class concentrations), were determined in livers of patients histologically classified as: non-NAFLD (n=21), simple steatosis (NAFL, n=44), steatohepatitis without significant fibrosis, stage of fibrosis F0-F1 (NASH, n=15) and steatohepatitis + stage of fibrosis F2-F4 (NASH-fibrosis, n=14). The values of Log_2_-fold change of each patient (column) and each lipid specie (row) is plotted in the in the heatmap and ordered by the hierarchical clustering of individuals (columns). (B) Hepatic concentrations of representative species of TG, dhSM, dhCer, dhHexCer, Cer and SM. Average values are represented as boxplots, with points showing the value of each patient. (C) Correlation plot between plasma and liver concentrations for Cer, dhCer and TG. *p <0.05, *p <0.01, ***p <0.001 and ****p <0.001, using Student’s t-test or the Pearson’s correlation coefficient. TG, triglyceride; CE, Cholesteryl ester; FC, free cholesterol; ACer, 1-o-Acylceramide; Cer, ceramide; HexCer, hexosylceramide; SM, Sphingomyelin; dhCer, dihydroceramide; dhHexCer, dihydrohexosylceramide dhSM, dihydrosphingomyelin. NASF; NASH-fibrosis

### Plasma lipidomic analysis of NAFLD patients

Previous liver biopsies were obtained from a cohort of obese patients with short representation of stages F3–F4, which did not allow comprehensive evaluation of the behaviour of lipid species in advanced fibrosis. Based on the correlation observed for dihydrosphingolipid species between liver and plasma, we analyzed plasma samples including those from patients with advanced fibrosis, with a view to defining the potential of plasma measurements as surrogate biomarkers of histological lesions. The clinical, biochemical and histological characteristics of the plasma cohort are presented in **Suppl. Fig. 7B** and **Table 2**. The average age (51.7 ± 11.2 years) and BMI (41.3 ± 9.0 kg/m^2^) of the whole cohort was not markedly different from that of the bariatric surgery cohort, but the proportion of females was slightly lower (59%, p=0.043), age of NASH-fibrosis patients was higher, and BMI lower relative to the other groups. Rates of hyperlipidemia, hypertension, T2DM and the trends in glycemic parameters were similar to those described for the bariatric surgery cohort. Transaminase levels showed a slight increase with progression from non-NAFLD to NASH-fibrosis. In the latter group, fibrogenesis test scores were significantly higher compared to the remaining groups (**Table 2**).

**Table 2.** Clinical and biochemical values in NAFLD patients from the plasma lipidomics cohort. Patients were classified according to their liver histological findings into four groups: non-NAFLD, simple steatosis (NAFL), steatohepatitis without significant fibrosis, stage of fibrosis F0-F1 (NASH) and steatohepatitis + stage of fibrosis F2-F4 (NASH-fibrosis). Values are expressed in % (N/N total in group) or mean ± SD or (N total in group). (a) Significant, p<0.05 from non-NAFLD. (b) Significant, p<0.05 from NAFL. (c) Significant, p<0.05 from NASH. (d) Significant, p<0.05 from NASH-fibrosis.

| Variable | non-NAFLD | NAFL | NASH | NASH-fibrosis |
| --- | --- | --- | --- | --- |
| **Demographics** |  |  |  |  |
| Number, % | 15 (29/195) | 33 (65/195) | 14 (27/195) | 38 (74/195) |
| Female, % | 69 (20/29) | 65 (42/65) | 63 (17/27) | 49 (36/74) |
| Obesity, % | 100 (29/29) | 96 (26/65) | 91 (59/27) | 76 (56/74) |
| Hypertension, % | 31 (9/29) | 46 (30/65) | 41 (11/27) | 70 (52/74) |
| T2DM, % | 21 (6/29) | 38 (25/65) | 44 (12/27) | 78 (58/74) |
| Dyslipidaemia, % | 31 (9/29) | 71 (46/65) | 67 (18/27) | 81 (60/74) |
| Bariatric Surgery, % | 100 (29/29) | 78 (51/65) | 63 (17/27) | 19 (14/74) |
| **Anthropometry** |  |  |  |  |
| Age, years | 42.3 ± 1.7 (29)<sup>b,c,d</sup> | 49.3 ± 1.1 (65)<sup>a,d</sup> | 50.0 ± 1.7 (27)<sup>a,d</sup> | 58.0 ± 1.3 (74)<sup>a,b,c</sup> |
| Weight, Kg | 127.6 ± 4.6 (29)<sup>c,d</sup> | 119.3 ± 3.0 (65)<sup>d</sup> | 115.1 ± 4.6 (27)<sup>a</sup> | 107.2 ± 3.8 (74)<sup>a,b</sup> |
| BMI, Kg/m<sup>2</sup> | 45.4 ± 1.3 (29)<sup>c,d</sup> | 43.4 ± 1.1 (65)<sup>d</sup> | 41.6 ± 1.5 (27)<sup>a,d</sup> | 37.7 ± 1.1 (74)<sup>a,b,c</sup> |
| **Laboratory tests** |  |  |  |  |
| Glucose, mg/dL | 93.6 ± 3.0 (28)<sup>b,c,d</sup> | 109.4 ± 4.9 (64)<sup>a,d</sup> | 120.1 ± 9.9 (27)<sup>a,d</sup> | 142.4 ± 6.2 (74)<sup>a,b,c</sup> |
| HbA1c, | 5.4 ± 0.1 (26)<sup>c,d</sup> | 6.0 ± 0.1 (61)<sup>d</sup> | 6.4 ± 0.3 (26)<sup>a,d</sup> | 7.0 ± 0.2 (67)<sup>a,b,c</sup> |
| HOMA-IR | 2.7 ± 0.4 (28)<sup>b,c,d</sup> | 4.1 ± 0.3 (58)<sup>a,d</sup> | 5.4 ± 0.7 (22)<sup>a,d</sup> | 10.4 ± 1.4 (43)<sup>a,b,c</sup> |
| Insulin, μU/mL | 12.0 ± 1.6 (29)<sup>c,d</sup> | 15.2 ± 1.1 (58)<sup>d</sup> | 18.0 ± 1.9 (22)<sup>a,d</sup> | 28.2 ± 3.2 (43)<sup>a,b</sup> |
| ALT, IU/mL | 28.1 ± 4.8 (28)<sup>c,d</sup> | 29.6 ± 1.9 (62)<sup>c,d</sup> | 38.6 ± 3.8 (27)<sup>a,b,d</sup> | 53.2 ± 3.5 (74)<sup>a,b,c</sup> |
| AST, IU/mL | 22.8 ± 4.3 (27)<sup>d</sup> | 24.4 ± 1.4 (63)<sup>d</sup> | 29.3 ± 2.7 (27)<sup>d</sup> | 43.0 ± 2.4 (74)<sup>a,b,c</sup> |
| GGT, IU/mL | 24.5 ± 4.9 (28)<sup>c,d</sup> | 32.1 ± 3.7 (62)<sup>c,d</sup> | 53.1 ± 9.2 (27)<sup>a,b,d</sup> | 104.4 ± 11.2 (74)<sup>a,b,c</sup> |
| Triglycerides, mg/dL | 107.3 ± 7.5 (27)<sup>b,c,d</sup> | 159.3 ± 11.2 (62)<sup>a</sup> | 157.9 ± 13.2 (27)<sup>a</sup> | 177.9 ± 12.0 (74)<sup>a</sup> |
| Cholesterol, mg/dL | 167.9 ± 6.5 (28) | 176.8 ± 4.9 (63) | 180.7 ± 9.1 (27) | 178.0 ± 4.1 (73) |
| HDLc, mg/dL | 45.0 ± 1.6 (28)<sup>d</sup> | 43.3 ± 1.4 (61) | 41.3 ± 1.9(27) | 41.3 ± 1.0 (74)<sup>a</sup> |
| LDLc, mg/dL | 105.0 ± 5.8 (26) | 106.2 ± 4.7 (61) | 110.9 ± 7.9 (27) | 106.7 ± 3.5 (73) |
| Albumin, g/dL | 3.9 ± 0.1 (29) | 3.9 ± 0.1 (65) | 4.0 ± 0.1 (27) | 3.9 ± 0.1 (74) |
| Plaq. counts, $\times 10^9$/L | 280.1 ± 11.1 (29)<sup>d</sup> | 264.2 ± 9.8 (64)<sup>d</sup> | 263.5 ± 14.5 (26)<sup>d</sup> | 190.6 ± 7.4 (74)<sup>a,b,c</sup> |
| **Fibrogenesis tests** |  |  |  |  |
| HA, ng/mL | 26.8 ± 4.0 (20)<sup>b,d</sup> | 38.0 ± 4.3 (42)<sup>a</sup> | 38.9 ± 6.2 (17)<sup>a,d</sup> | 135.6 ± 27.0 (37)<sup>a,b,c</sup> |
| PIIINP, ng/mL | 5.5 ± 0.4 (20)<sup>b,c,d</sup> | 7.0 ± 0.4 (42)<sup>a</sup> | 7.8 ± 1.2 (17)<sup>a,d</sup> | 11.2 ± 0.9 (37)<sup>a,b,c</sup> |
| TIMP, ng/mL 1 | 122.9 ± 6.6 (20)<sup>b,c,d</sup> | 168.4 ± 9.3 (42)<sup>a</sup> | 192.4 ± 15.3(17)<sup>a,d</sup> | 268.3 ± 17.2 (37)<sup>a,b,c</sup> |
| CK18, U/L | 117.9 ± 16.1 (21)<sup>d</sup> | 177.2 ± 29.8 (42)<sup>d</sup> | 176.6 ± 35.5 (17)<sup>d</sup> | 331.6 ± 32.0 (36)<sup>a,b,c</sup> |
| ELF score | 8.0 ± 0.2 (20)<sup>b,c,d</sup> | 8.6 ± 0.1 (42)<sup>a,d</sup> | 8.7 ± 0.2 (17)<sup>a,d</sup> | 10.0 ± 0.2 (37)<sup>a,b,c</sup> |
| FIB-4 score | 0.649 ± 0.062 (27)<sup>b,c,d</sup> | 0.939 ± 0.061 (60)<sup>a</sup> | 0.979 ± 0.116 (26)<sup>a,d</sup> | 2.156 ± 0.154 (74)<sup>a,b,c</sup> |
| NFS score | - 0.973 ± 0.236 (27)<sup>d</sup> | - 0.466 ± 0.209 (60)<sup>d</sup> | - 0.637 ± 0.306 (26)<sup>d</sup> | 0.700 ± 0.144 (74)<sup>a,b,c</sup> |
| ***Plasma lipid classes*** |  |  |  |  |
| TG, nmol/mL | 233.0 ± 13.8 (29)<sup>b,c,d</sup> | 297.7 ± 10.3 (65)<sup>a</sup> | 307.4 ± 20.5 (27)<sup>a</sup> | 318.1 ± 13.2 (74)<sup>a</sup> |
| CE, nmol/mL | 3574 ± 272 (29)<sup>b</sup> | 4254 ± 191 (65)<sup>a,d</sup> | 4097 ± 303 (27) | 3634 ± 143 (74)<sup>b</sup> |
| FC, nmol/mL | 1370 ± 50 (29)<sup>b</sup> | 1554 ± 44 (65)<sup>a</sup> | 1513 ± 78 (27) | 1460 ± 35 (74) |
| ACer, nmol/mL | 0.153 ± 0.009 (29)<sup>b,c,d</sup> | 0.194 ± 0.007 (65)<sup>a</sup> | 0.192 ± 0.013 (27)<sup>a,d</sup> | 0.229 ± 0.01 (74)<sup>a,b,c</sup> |
| Cer, nmol/mL | 5.5 ± 0.3 (29)<sup>b,d</sup> | 6.5 ± 0.2 (65)<sup>a</sup> | 6.3 ± 0.4 (27) | 6.3 ± 0.2 (74)<sup>a</sup> |
| HexCer, nmol/mL | 6.8 ± 0.4 (29) | 7.0 ± 0.3 (65) | 7.3 ± 0.5 (27) | 7.5 ± 0.3 (74) |
| SM, nmol/mL | 355.3 ± 13.2 (29) | 372.6 ± 8.7 (65)<sup>d</sup> | 388.4 ± 21.5 (27) | 349.4 ± 7.7 (74)<sup>b</sup> |
| dhCer, nmol/mL | 0.66 ± 0.05 (29)<sup>d</sup> | 0.76 ± 0.03 (65)<sup>d</sup> | 0.76 ± 0.06 (27) | 0.87 ± 0.04 (74)<sup>a,b</sup> |
| dhHexCer, nmol/mL | 0.044 ± 0.002 (29)<sup>d</sup> | 0.049 ± 0.002 (65)<sup>d</sup> | 0.051 ± 0.004 (27)<sup>d</sup> | 0.061 ± 0.003 (74)<sup>a,b,c</sup> |
| dhSM, nmol/mL | 33.7 ± 1.5 (29) | 32.9 ± 1.2 (65) | 33.1 ± 1.8 (27) | 33.8 ± 1.3 (74) |
| LPC, nmol/mL | 131.1 ± 8.0 (29)<sup>b,d</sup> | 153.3 ± 6.8 (65)<sup>a</sup> | 154.2 ± 9.2 (27) | 175.4 ± 6.8 (74)<sup>a,b</sup> |
| PC, nmol/mL | 1071 ± 45 (29)<sup>b,d</sup> | 1192 ± 39 (65)<sup>a</sup> | 1193 ± 59 (27) | 1249 ± 44 (74)<sup>a</sup> |
| PE, nmol/mL | 41.5 ± 3.4 (29)<sup>b,c,d</sup> | 51.4 ± 2.3 (65)<sup>a</sup> | 57.0 ± 4.6 (27)<sup>a</sup> | 57.0 ± 3.1 (74)<sup>a</sup> |

T2DM. Type II diabetes mellitus ; HbA1c, Glycated haemoglobin; HOMA-IR, homeostatic model assessment for insulin resistance; ALT, alanine aminotransferase; AST, aspartate aminotransferase; GGT, Gamma glutamyltransferase; BMI, body mass index; HDLc, high-density lipoprotein cholesterol; LDLc, Low-density lipoprotein cholesterol; HA, Hyaluronic Acid; PIIINP, Procollagen III aminoterminal peptide; TIMP1, Tissue inhibitor of metalloproteinase 1; CK18, Apoptosis-associated caspase-cleaved keratin 18. ELF, Enhanced liver fibrosis score; FIB-4, Fibrosis-4 index; NFS, NAFLD fibrosis score.

Overall, lipidomic analysis of plasma samples of the full cohort revealed higher neutral lipid and dihydrosphingolipid (dhCer, dhSM and dhHexCer) concentrations in NAFLD compared to non-NAFLD patients (**Fig. 6A, B**). Similar to the findings in liver of both patients and model mice, Cer and SM levels were not significantly affected by disease progression. However, we did not detect an increase in plasma dihydrosphingolipid in patients with advanced fibrosis stages and lower steatosis scores, similar to data obtained with mice, reflecting that plasma lipid changes clearly do not represent a perfect surrogate of changes in liver.

**Fig. 6.**
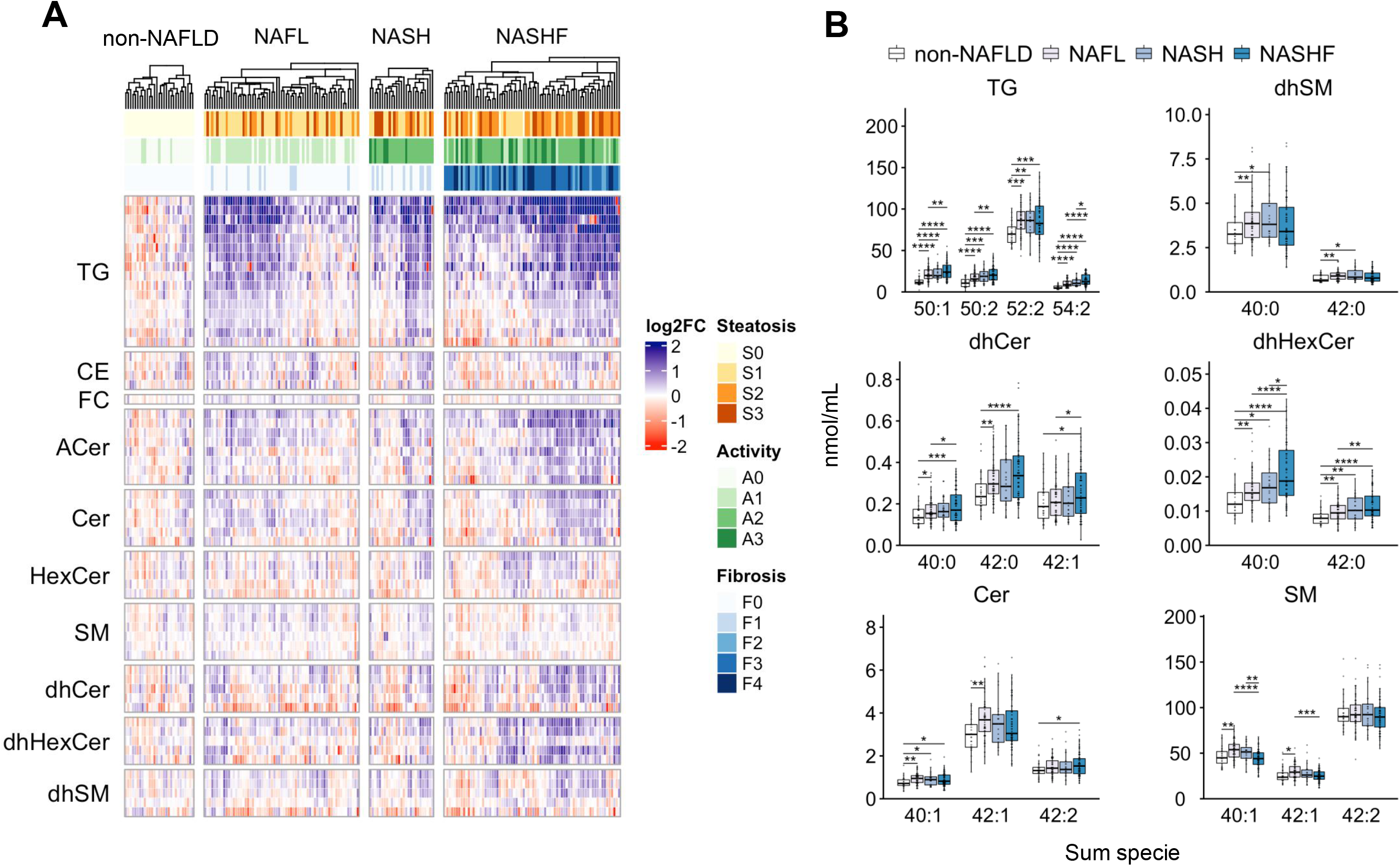
Lipidomic analysis of (dihydro)sphingolipids in plasma of NAFLD patients. (A) Quantitative analysis of lipid species from 10 classes (see Table 2 for lipid class concentrations), were determined in livers of patients histologically classified as: non-NAFLD (n=29), simple steatosis (NAFL, n=65), steatohepatitis without significant fibrosis, stage of fibrosis F0-F1 (NASH, n=27) and steatohepatitis + stage of fibrosis F2-F4 (NASH-fibrosis, n=74). The values of Log_2_-fold change of each patient (column) and each lipid specie (row) is plotted in the in the heatmap and ordered by the hierarchical clustering of individuals (columns). (B) Hepatic concentrations of representative species of TG, dhSM, dhCer, dhHexCer, Cer and SM. Average values are represented as boxplots, with points showing the value of each patient. *p <0.05, *p <0.01, ***p <0.001 and ****p <0.001, using Student’s t-test. TG, triglyceride; CE, Cholesteryl ester; FC, free cholesterol; ACer, 1-o-Acylceramide; Cer, ceramide; HexCer, hexosylceramide; SM, Sphingomyelin; dhCer, dihydroceramide; dhHexCer, dihydrohexosylceramide dhSM, dihydrosphingomyelin. NASF; NASH-fibrosis

## DISCUSSION

NAFLD is a complex multifactorial disease with a histological spectrum spanning from generally benign steatosis (NAFL) to steatosis with evidence of hepatocellular inflammation and damage (NASH). The latter symptom appears in about 25% NAFLD patients, among which 5% go on to develop NASH-fibrosis (26). The available evidence supports that overnutrition causing central obesity and insulin resistance defines metabolic predisposition to NASH (27). Ceramides and their immediate precursors, dihydroceramides, are implicated in NAFLD and other comorbidities, such as obesity and T2DM (6). However, the relationship of (dihydro)sphingolipid species with NAFLD progression remains to be established.

Here, we firstly explored the role of (dihydro)sphingolipids in a mouse model of NAFLD. To date, no animal models have fully reproduced all the metabolic and histological hallmarks of human NAFLD (28, 29). A minimum set of requirements in the animal model is demonstration of metabolic aspects related to disease such as obesity, IR, and dyslipidemia, in addition to progressive evolution of liver damage (26). Our regime of choice consisted of a high-fat and cholesterol diet supplemented with high fructose and glucose. Glucose promotes hyperinsulinemia, which activates liver FA synthesis by increasing mRNA expression and proteolytic cleavage of the nuclear transcription factor SREBP1c (30), while fructose acts synergistically to promote *de novo* lipogenesis and block β-fatty acid oxidation, leading to NAFLD development (31). Addition of high cholesterol (1.25%) accelerates NASH development, directly activating myofibroblast transformation of hepatic stellate cells (HSCs) (32). As a result, our model mice developed obesity and IR at 22 weeks accompanied by progressive transaminase elevation and hypercholesterolemia. However, hypertriglyceridemia was absent in our model, consistent with previous findings (33–35). The reason for this phenomenon is not completely clear but may be attributed to the differences in lipoprotein metabolism between humans and mice. The latter lacks cholesteryl ester transfer protein, which may reduce the secretion of TG containing very-low density lipoproteins (VLDL) by liver and stimulate high-density lipoprotein (HDL) production (36).

The model additionally demonstrated histological progression from NAFL to NASH, as evident from the presence of a cluster of inflammatory cells and ballooned hepatocytes at 40 weeks. The latter finding was confirmed by increased protein expression of p62, a proteasome component and marker of Mallory-Denk bodies commonly found in human NASH (37). HSC activation and myofibroblast transformation were validated by the increased protein and gene expression of α-SMA and *Tfgb1*, respectively. Immunohistochemical analysis revealed association of the α-SMA marker with formation of hCLS, linked with macrophage Kupffer cells surrounding cholesterol-loaded hepatocytes and synthesis of collagen (38, 39).

One disadvantage of our murine model was that even after long-term feeding, disease in animals did not progress to advanced fibrosis stages, consistent with earlier findings (40). For this reason, we combined HFD feeding with CCl_4_ as a fibrosis accelerator as reported in previous studies (33). We observed progression to stages F3-F4 in 40% (3 out of 5) mice at 6 weeks and 100% animals after 10 weeks of CCl_4_ treatment (33). However, CCl_4_ administration was associated with accelerated weight loss and amelioration of IR. Attenuation of IR could result from the blunted weight gain, but was also observed in the HFD group of mice at 40 weeks, suggestive of effects of fibrogenesis on IR. In summary, our mouse model could effectively replicate some of the key metabolic, inflammatory and profibrotic pathways of human NAFLD.

Lipidomic analyses showed that changes in dihydrosphingolipid species from liver and plasma in the mouse model were more significant than those in sphingolipids. Relative to sphingolipid species that are highly abundant in the diet, the dihydrospingolipid content is extremely low (41). The majority of dihydrosphingolipids were obtained directly from stimulation of *de novo* synthesis in response to FFA overload, in keeping with earlier reports (8), and confirmed by the increase in plasma and liver FFA concentrations in parallel with higher gene expression of *Cd36*, a scavenger receptor that mediates the binding and cellular uptake of FFA (11) and stearoyl-CoA desaturase-1 (*Scd1*), a direct target of SREBP1c, which stimulates fatty acid desaturation and TG synthesis (42). Moreover, the coordinated increase in gene expression of enzyme isoforms of *Sptlc1-2, Cers2-6, Degs1* and *Sgms1-2* supports that liver of NAFLD mouse responds to the FFA surplus, triggering not only synthesis of neutral lipids (TG, CE) but also early activation of the ceramide synthesis pathway. In this context, increase in 1-o-acylceramide (ACer) catalyzed by the same diacylglycerol acyltransferase 2 (DGAT2) responsible for TG synthesis (43), albeit at levels four orders in magnitude lower in quantitative terms compared with TG, could be interpreted as a result of the same spillover mechanism induced by excess liver FFAs.

Changes in neutral lipid classes (represented by TG, CE and ACer) were correlated with histology and reflected in the steatosis score. However, advanced fibrosis stages (F3-F4) were associated with lower steatosis scores and contents of neutral lipids. This phenomenon is similar to the reduction of hepatic fat observed in patients with cryptogenic cirrhosis of NAFLD etiology (burnt-out NASH) (44). However, advanced fibrosis dissociates the increase in dihydrosphingolipid from the steatosis-linked neutral lipid increase observed in the mouse model (see **Suppl. Fig 5**). We speculate that advanced fibrosis triggers the expression of sphingomyelinases (45), ceramidases (46) and sphingosine kinases (47), reducing the available pool of Cer, in turn, promoting *de novo* sphingolipid synthesis and increasing the dihydrosphingolipid concentration.

The concentrations of dihydrosphingolipids in mouse liver increased from NAFL to NASH-fibrosis, especially those with VLCFA, such as dhCer 40:0, dhCer 42:0 and dhCer 42:1, whereas Cer and SM-related species showed no changes or a decreasing trend with disease progression. The general view is that liver respond to HFD feeding triggering the accumulation of C16-Cer and decreasing the content of VLCFA-Cer (10, 48–50). However, more recently, loss of *Cers2* function, which partially blocked the synthesis of VLCFA-Cer, failed to increase the Cer 34:1 concentration in liver, even in mice fed a palmitic acid-enriched diet (51). The diet utilized in our study contained a higher proportion of cholesterol and lower palmitic acid content than reported previously (10, 48, 49). Therefore, we hypothesize that the high-cholesterol content of our diet is responsible for differences in the distribution of (dihydro)sphingolipid species relative to previous studies. The free cholesterol (FC) content was almost double in liver of our NALFD mice (**Suppl. Table S1**). A major proportion of the excess content is esterified as CE, but the increase in FC is likely to affect hepatocyte membrane composition where it is known to displace Cer, in particular, shorter C16-Cer, and associate preferentially with VLCFA-Cer, SM and dihydrosphingolipid species (52).

Histological findings in the livers of bariatric surgery patients and plasma of the full NAFLD cohort were similar to those in the mouse model upon classification according to SAF score (21). Moreover, metabolic characteristics, dyslipidemia, IR and T2DM, evolution of fibrogenesis biomarkers and even transaminase changes progressed in a comparable manner from non-NALFD to NASH-fibrosis. Lipidomic data from patient biopsies additionally showed many similarities in the trends observed for neutral lipids (TG, CE and ACer) with the mouse model. Interestingly, we observed equivalent changes in liver samples of NAFLD patients, with an increase in VLCFA-dihydrosphingolipid species and no changes in Cer and SM groups, in keeping with recent publications. On one hand, Vvedenskaya et al. (19) observed no major changes other than neutral TG, DG and CE contents in liver between NAFL and NASH patient groups. Notably, the concentrations of Cer and SM were similar between NAFL and NASH. On the other hand, Ooi and co-workers (18) reported changes in dihydrosphingolipid species between non-NAFLD and NAFLD but no differences between NAFL and NASH patient groups. Significant differences were additionally observed for VLCFA-dhCer between the latter two groups.

Our data from model mice and patients support the theory that the increase in dihydrosphingolipid is mediated by upregulation of *de novo* ceramide synthesis and linked to higher steatosis scores. We observed a good correlation of dhCer levels between liver and plasma and TG species in liver of patients but no significant associations for Cer species. Secretion of dhCer is associated with TG and VLDL production, as recently shown by Carlier et al.(17). The same authors reported that some dhCer species in plasma are associated with hepatic steatosis and NASH in T2DM patients as well as increased liver gene expression of *Sptlc1, Degs1, Sgms1,* similar to data obtained with our mouse model. Moreover, plasma dhCer was reported to be higher in individuals who eventually progress to diabetes up to 9 years before disease onset (53), in agreement with our interpretation of early stimulation of the *de novo* sphingolipid pathway in comorbidities directly associated with NAFLD.

In view of the existence of good correlations between dihydrosphingolipid concentrations in the liver and plasma, we were encouraged to extend the study to a larger cohort of patients with advanced fibrosis stages to determine the potential of measurement of dihydrosphingolipids in plasma as a biomarker of disease progression. Previous studies that attempted to address this question recruited limited patients with significant fibrosis (F2-F4) (16, 18, 19, 54). Certain species of TG (TG 50:1, TG 50:2, TG 52:2 and TG 54:2) in plasma appeared especially well suited to differentiate between non-NAFL and NAFLD, even when fibrosis was observed, in agreement with earlier findings. Plasma dihydrosphingolipids showed more significant associations with disease progression than sphingolipids but their performance was not much better than that of TG species. Overall, the benefit of measuring these lipid species over available low-cost laboratory assessments of parameters, such as transaminases, and validated fibrosis scores, such as NAFLD fibrosis score (NFS), Fibrosis-4 index (FIB-4) and Enhanced liver fibrosis score (ELF), appears to be of limited utility in stratification of patients with NAFL, NASH and NASH-fibrosis.

One of the major strengths of our study is the detailed descriptions of liver and plasma lipid profiles in the mouse model and NAFLD patient cohort, which showed similar histological and metabolic alterations. The concentrations of (dihydro)sphingolipids and other lipid species were analyzed following a validated methodology (24), which demonstrated excellent analytical stability. The results were expressed as a molar concentration and shared in an open repository, which facilitates future comparison with other lipidomic datasets from NAFLD patients and ongoing efforts to harmonize methodology (55).

However, our study has several limitations that should be taken into consideration. Although the transcriptomic and histological evolution of the mouse model is similar to human NAFLD (33), mice in our experiments were subjected to an extreme diet regime and fibrosis accelerated with CCl_4_, a toxic agent which has no implications in human pathology (56). We could not differentiate whether the metabolic findings in NASH-fibrosis mice were potentially driven by weight loss induced by fibrosis evolution or whether progressive fibrosis was responsible for weight loss. In fact, advanced fibrosis is associated with a hypermetabolic state (57), which may explain the decrease in weight observed at the NASH-fibrosis stage in the mouse model and patients (see **Table 2**). We hypothesized that similar changes in (dihydro)sphingolipid species to those observed in the liver of mice are likely to occur in patients with NASH-fibrosis. Unfortunately, we could only analyze the concentrations of lipid species in liver samples of six patients with advanced fibrosis from the bariatric surgery cohort. Hence, due to the inherent restrictions imposed by low availability of samples obtained in liver percutaneous biopsies, we were only able to analyze plasma samples from the cohort of patients with advanced fibrosis. Our results showed that although significant, at best, changes in dihydrosphingolipid species observed in plasma of NAFLD patients explained no more than *∼*50% of those in the liver (R=0.52, see **Fig. 5**). Therefore, we could not fully extrapolate the trends in plasma to those in liver of patients. Moreover, we did not evaluate the effects of the several genetic polymorphisms directly related to lipid metabolism, which are implicated in changes observed in liver ceramides (15). Nevertheless, earlier studies indicate that these risk genes do not seem to induce alterations in the liver lipidome (19). Another drawback, is that non-NAFLD patient liver and plasma samples were obtained from obese individuals without histologically proven disease and not truly healthy subjects and it might well be that (dihydro)sphingolipid concentrations were higher in non-NAFLD obese than in healthy individuals. Besides, the prospective single-point sampling nature of the study in both bariatric surgery and plasma cohorts of patients did not allow us to draw conclusions about the stability of performed measurements over time. Due to the inherent complexity of NAFLD, coordinated efforts including studies on larger sets of NAFLD patients with histologically defined grading and staging of the disease, elucidation of metabolic conditions and genetic risks, along with research focus on harmonization of lipidomic analytical approaches are essential to define the roles of lipid species in disease pathophysiology (58).

In conclusion, dihydrosphingolipid species accumulate in the liver of NAFLD patients, coincident with the patterns observed in a mouse model. The increase in dihydrosphingolipids reflects early stimulation of the *de novo* ceramide pathway in response to an increased burden of free fatty acids in the liver. The concentrations of dihydrosphingolipids, particularly those with very-long-chain fatty acids, correlates with liver triglycerides and steatosis grading based on histology. Interestingly, the appearance of advanced fibrosis downregulates accumulation of neutral lipids, but not dihydrosphingolipids in liver in the mouse model. While plasma dihydrosphingolipids do not appear to achieve a higher diagnostic accuracy than the currently available clinical chemistry tests, they provide valuable insights into the roles of lipid species in NAFLD progression. Further studies combining standardized quantitative lipidomic and proteomic approaches in well-defined cohorts of NAFLD patients are warranted to resolve the unmet need for plasma surrogate biomarkers of histological lesions in these patient populations.

## Data Availability

All data produced in the present study are available upon reasonable request to the authors

https://doi.org/10.5281/zenodo.6122303

## AKNOWLEDGMENTS

We thank the Quantification and Molecular Characterization Unit (UCA-CCM IRYCIS) for their technical help. JL Rodríguez-Navarro for helping with animals. P. Mediavilla, D. Mora, L. Alcázar and I. Candela for their assistance with blood biochemistry assays and International Science Editing for English style revision.

---

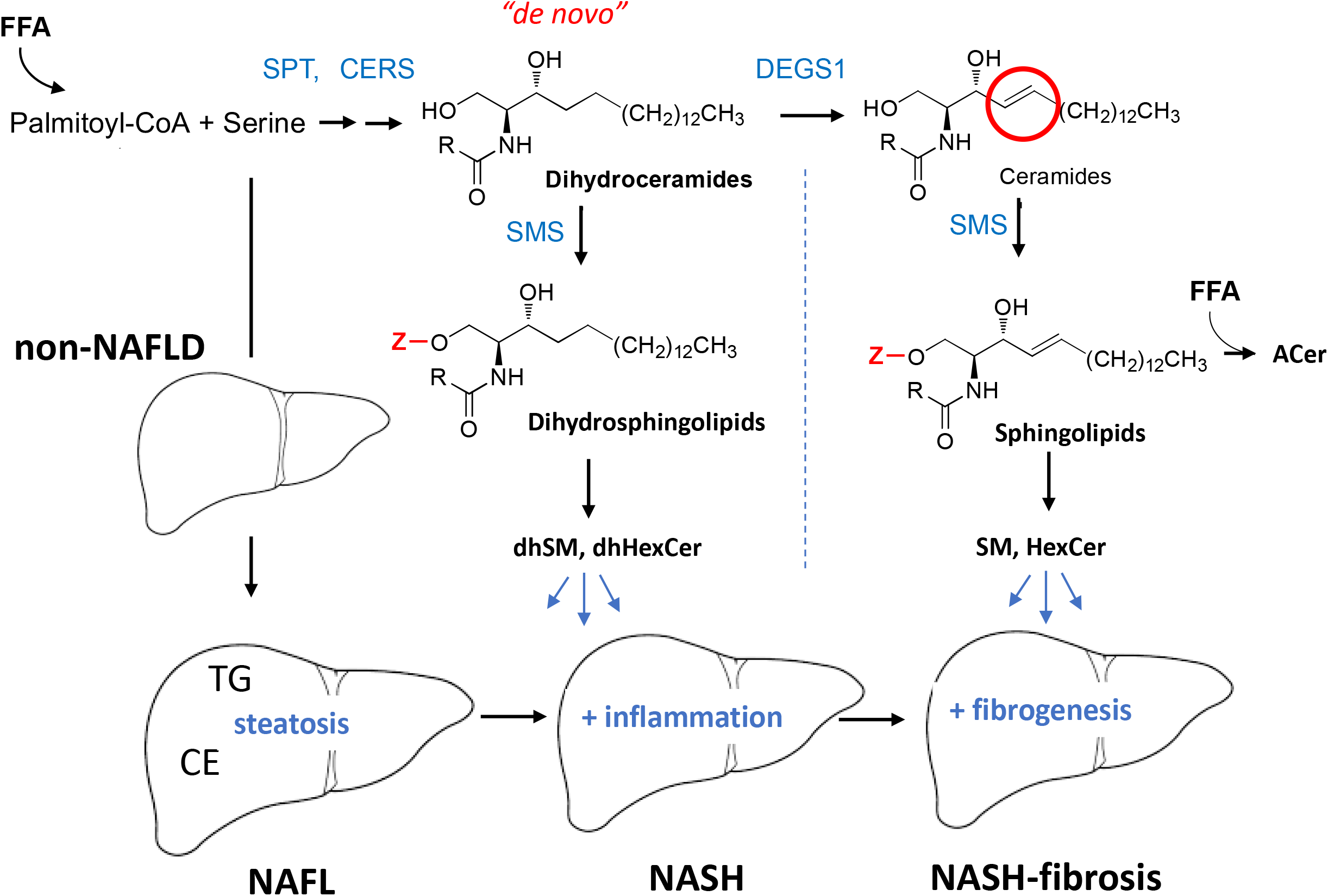

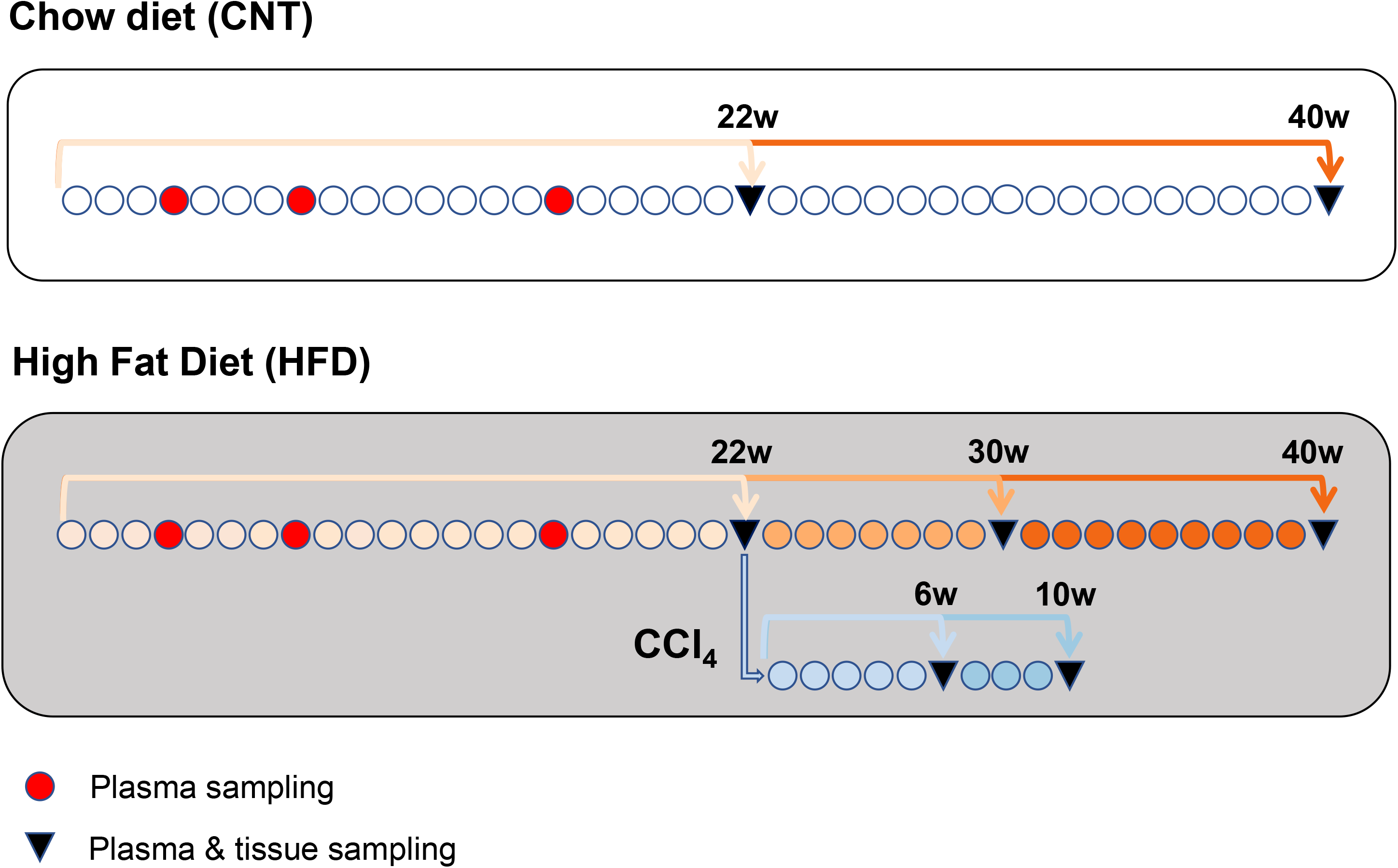

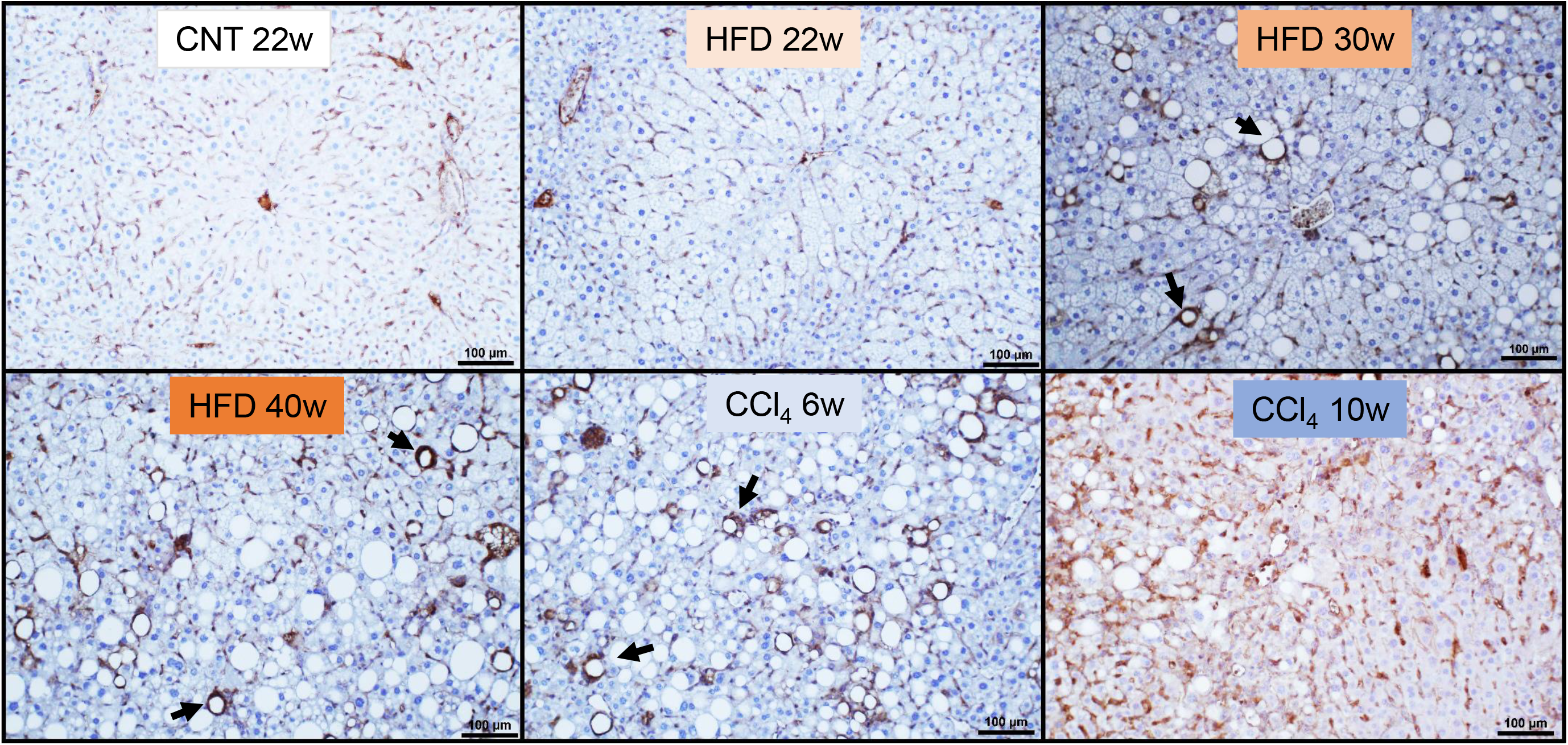

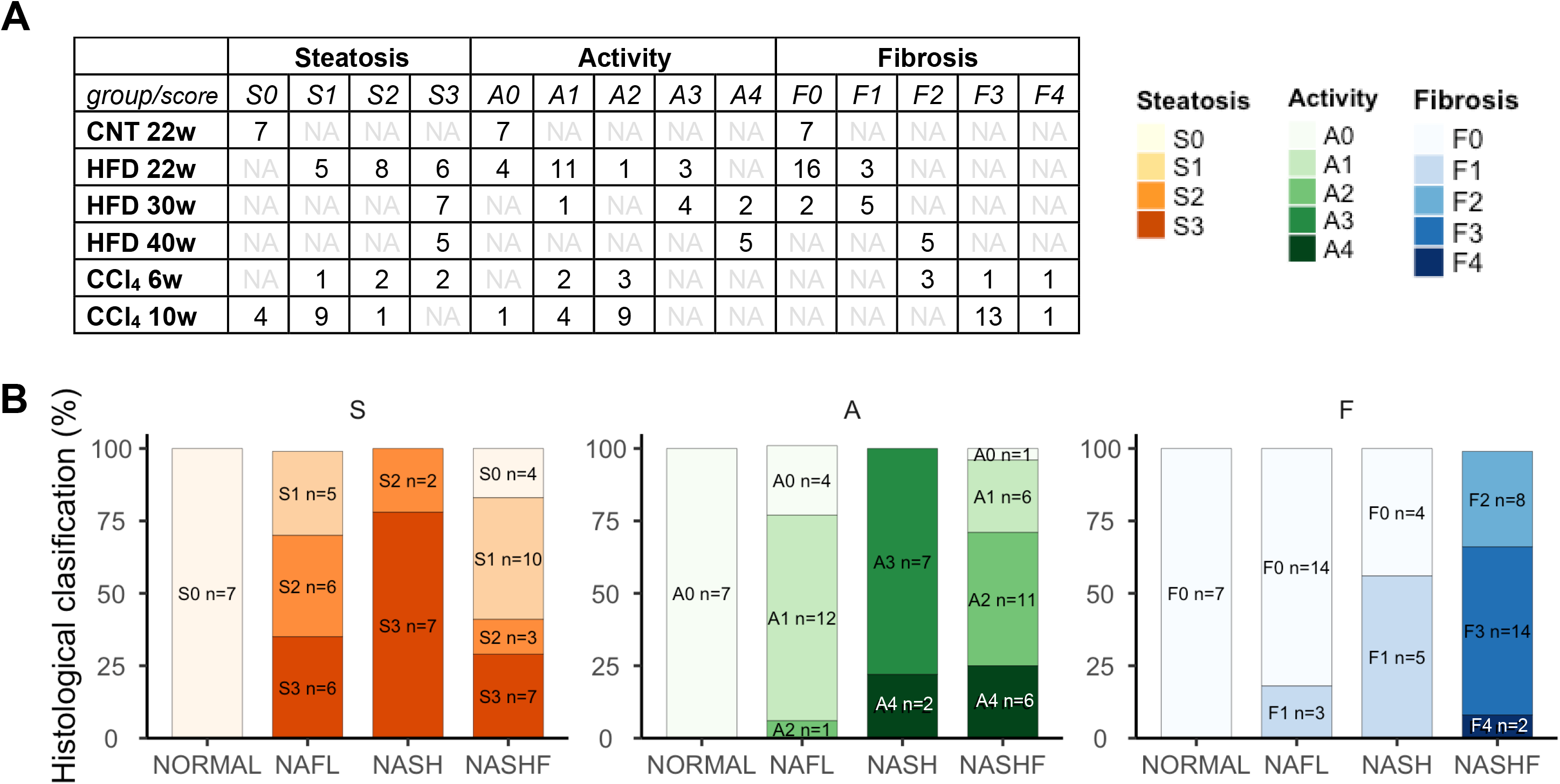

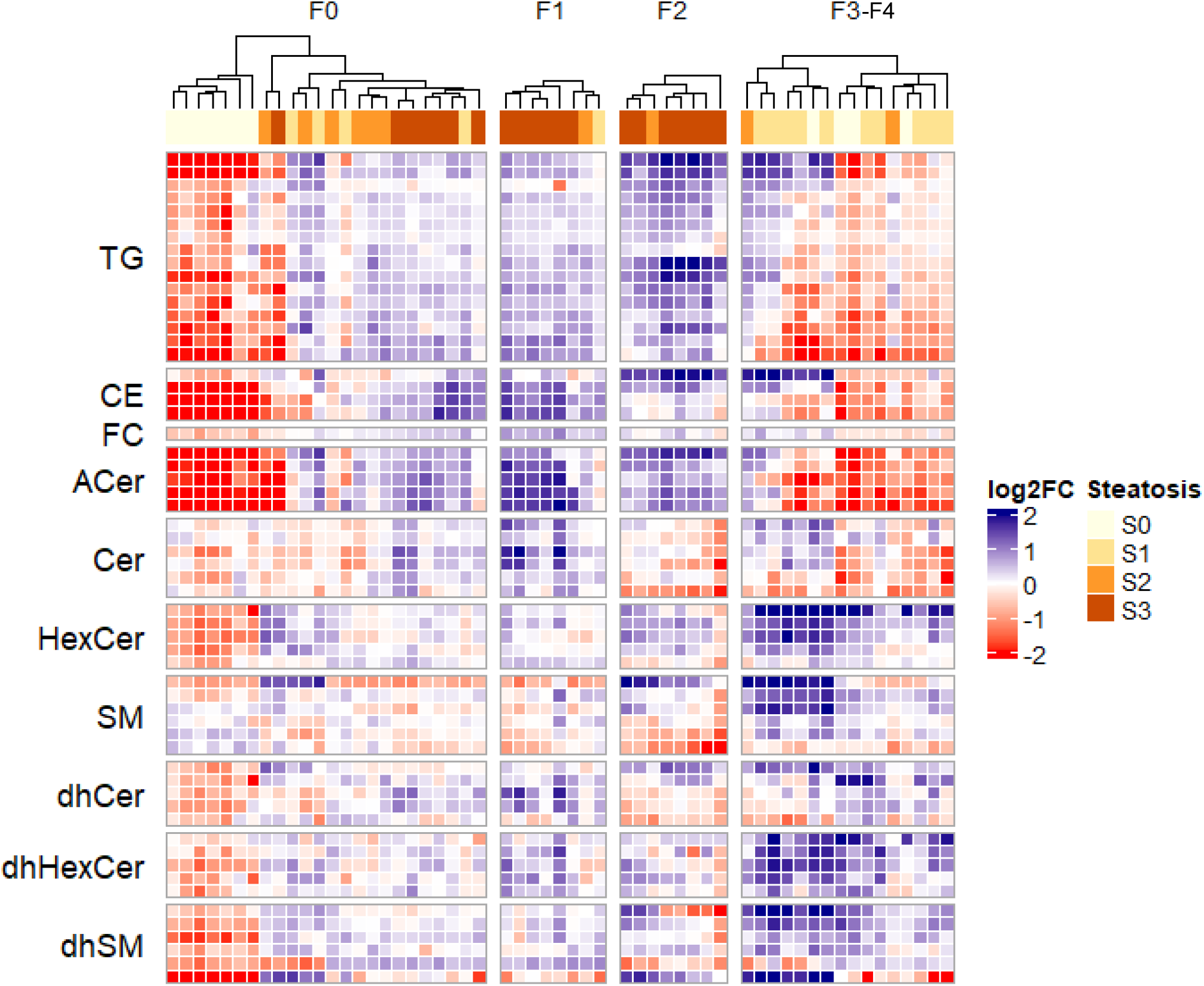

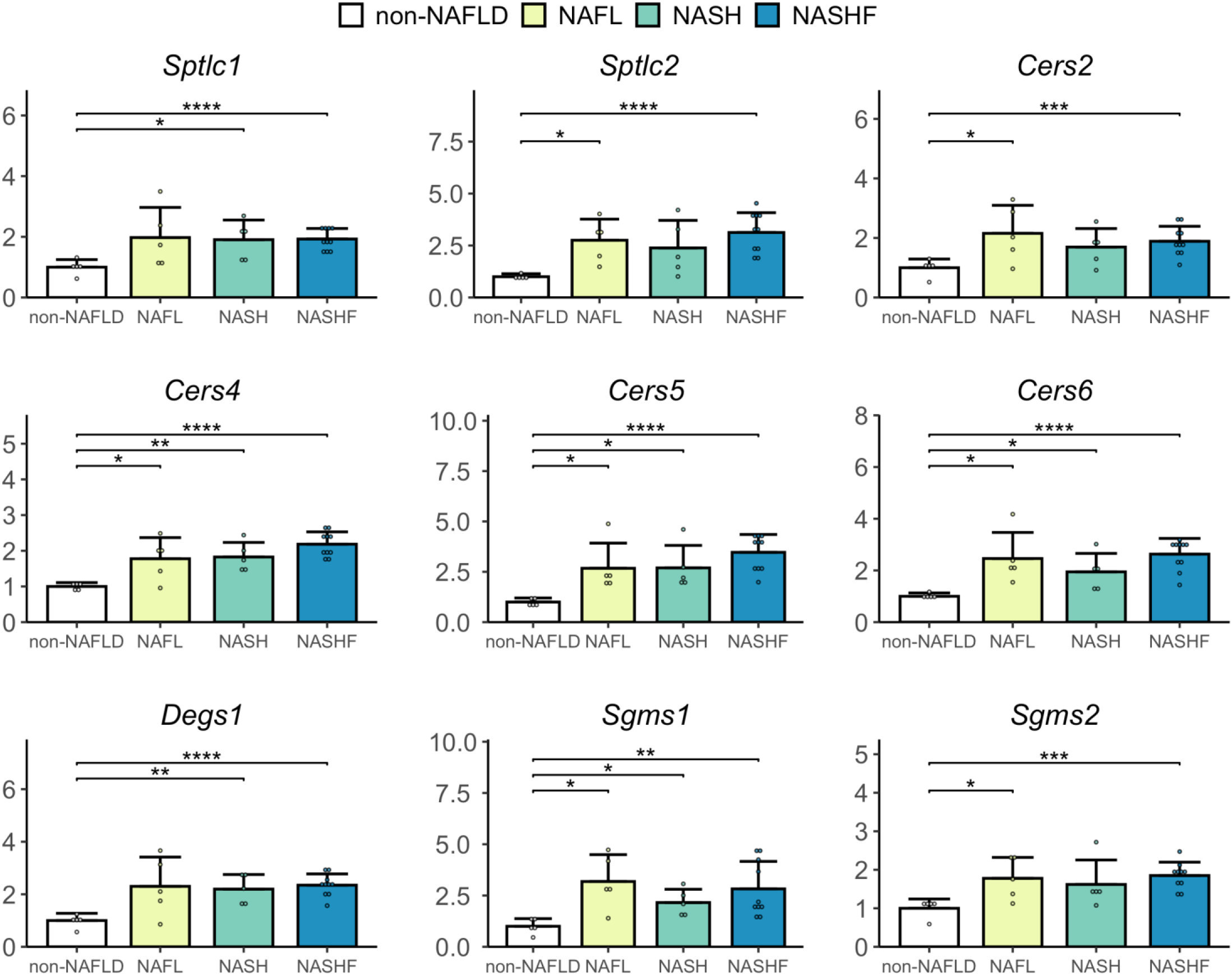

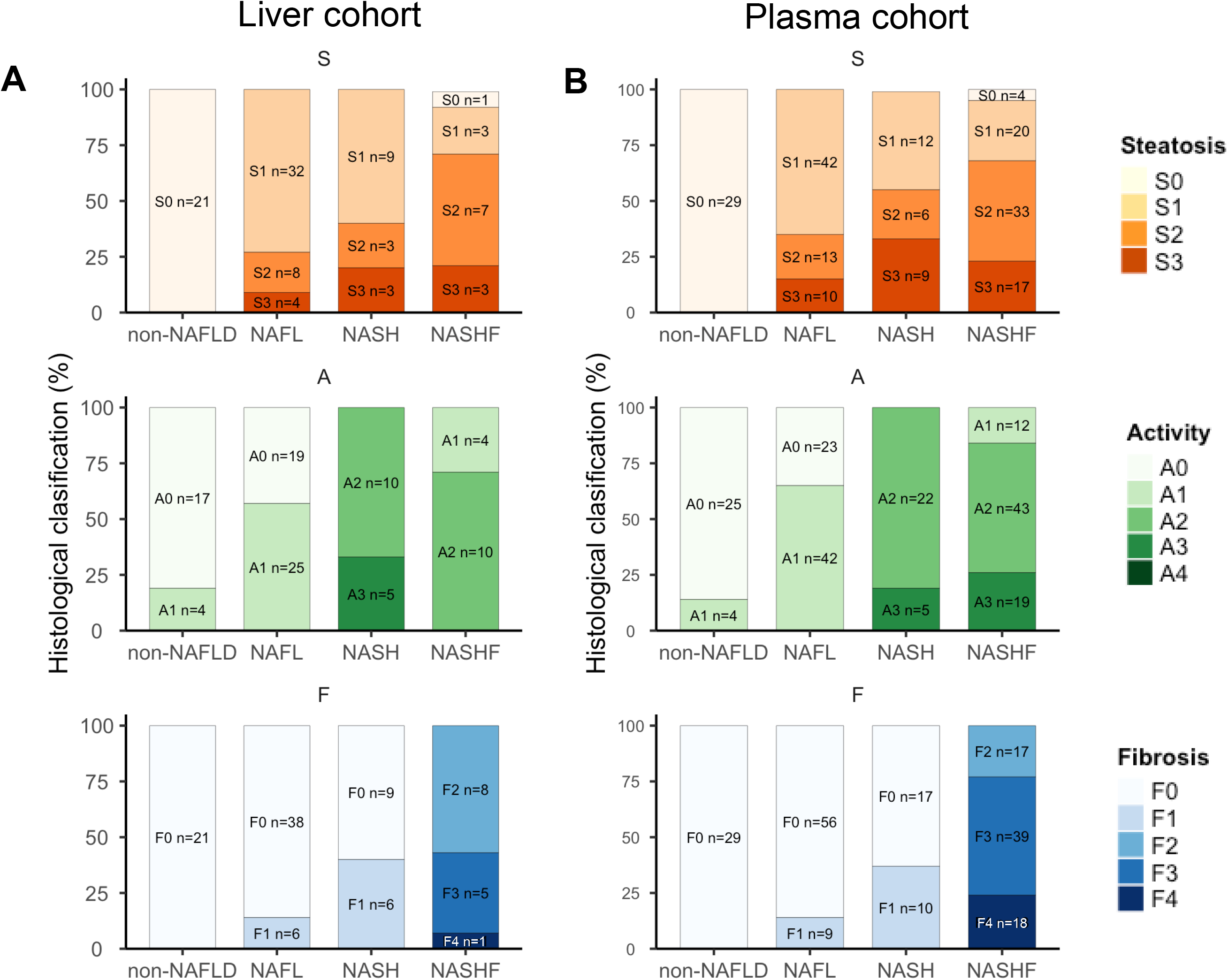

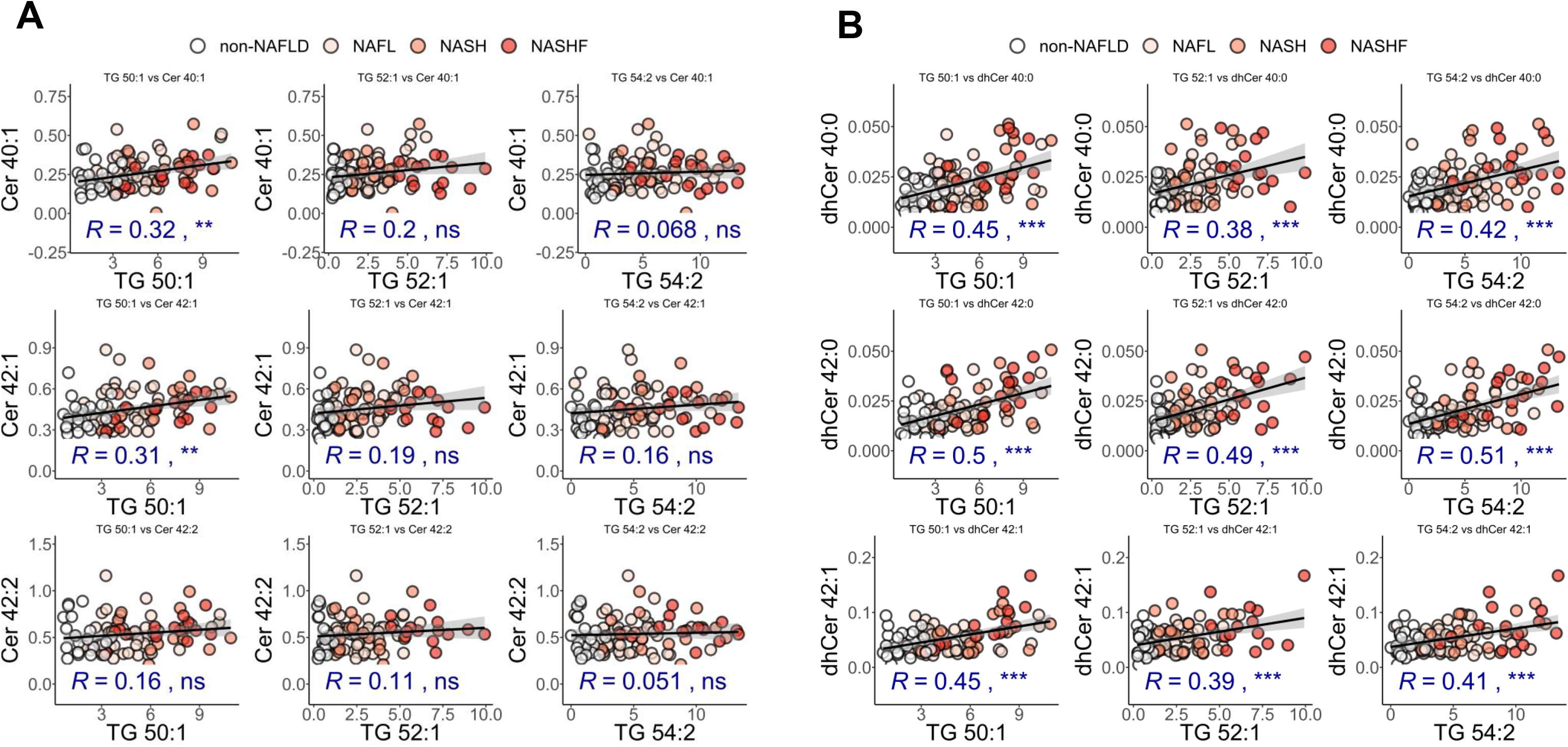

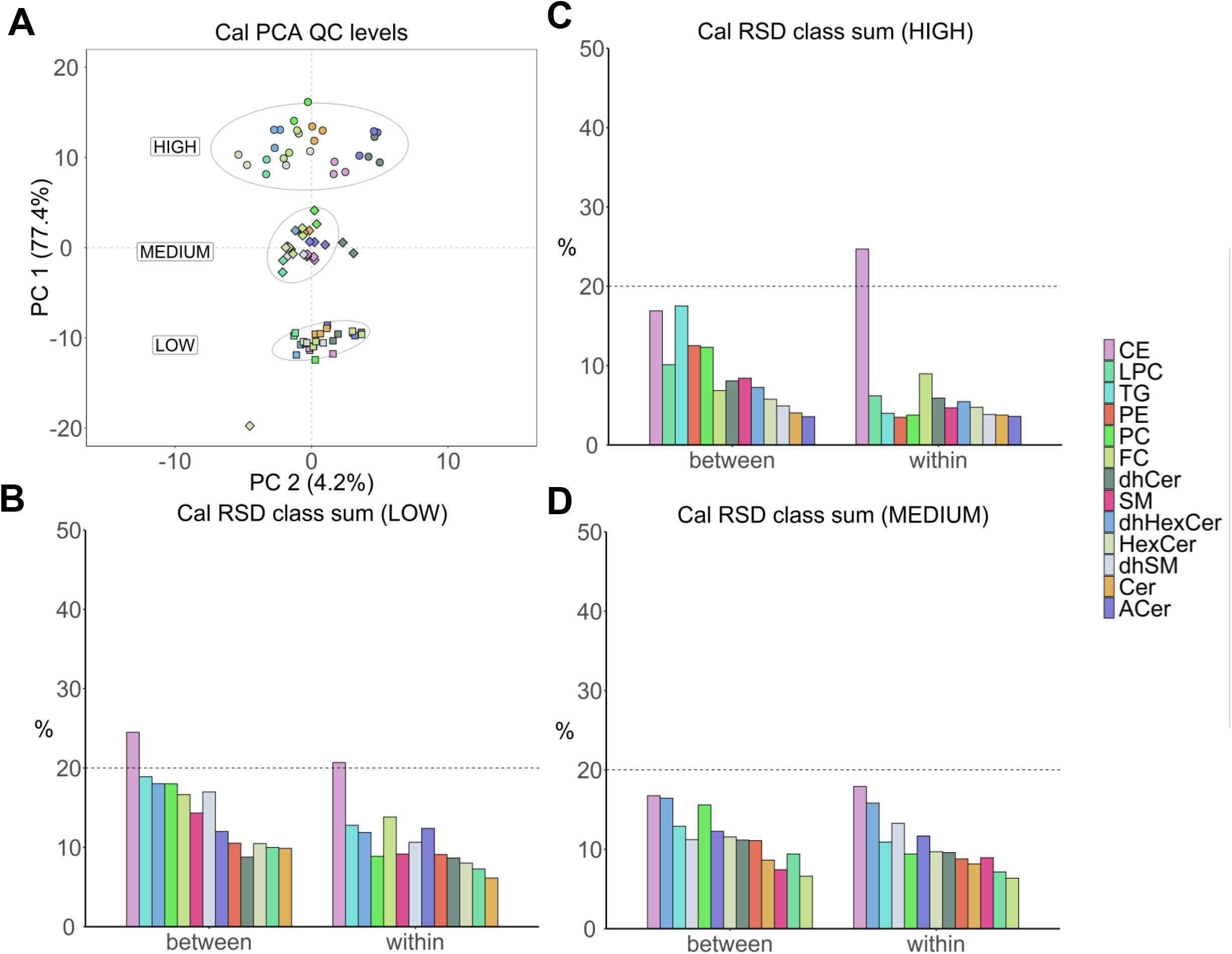

## REFERENCES

1. Eslam M, Newsome PN, Sarin SK, Anstee QM, Targher G, Romero-Gomez M, et al. A new definition for metabolic dysfunction-associated fatty liver disease: An international expert consensus statement. Journal of hepatology. 2020;73(1):202–9.

2. Chalasani N, Younossi Z, Lavine JE, Charlton M, Cusi K, Rinella M, et al. The diagnosis and management of nonalcoholic fatty liver disease: Practice guidance from the American Association for the Study of Liver Diseases. Hepatology (Baltimore, Md). 2018;67(1):328–57.

3. Angulo P, Kleiner DE, Dam-Larsen S, Adams LA, Bjornsson ES, Charatcharoenwitthaya P, et al. Liver Fibrosis, but No Other Histologic Features, Is Associated With Long-term Outcomes of Patients With Nonalcoholic Fatty Liver Disease. Gastroenterology. 2015;149(2):389–97 e10.

4. Sanyal AJ, Van Natta ML, Clark J, Neuschwander-Tetri BA, Diehl A, Dasarathy S, et al. Prospective Study of Outcomes in Adults with Nonalcoholic Fatty Liver Disease. The New England journal of medicine. 2021;385(17):1559–69.

5. Samuel VT, Shulman GI. Nonalcoholic Fatty Liver Disease, Insulin Resistance, and Ceramides. The New England journal of medicine. 2019;381(19):1866–9.

6. Hajduch E, Lachkar F, Ferre P, Foufelle F. Roles of Ceramides in Non-Alcoholic Fatty Liver Disease. J Clin Med. 2021;10(4).

7. Green CD, Maceyka M, Cowart LA, Spiegel S. Sphingolipids in metabolic disease: The good, the bad, and the unknown. Cell metabolism. 2021;33(7):1293–306.

8. Summers SA, Chaurasia B, Holland WL. Metabolic Messengers: ceramides. Nat Metab. 2019;1(11):1051–8.

9. Jiang M, Li C, Liu Q, Wang A, Lei M. Inhibiting Ceramide Synthesis Attenuates Hepatic Steatosis and Fibrosis in Rats With Non-alcoholic Fatty Liver Disease. Front Endocrinol (Lausanne). 2019;10:665.

10. Kim YR, Lee EJ, Shin KO, Kim MH, Pewzner-Jung Y, Lee YM, et al. Hepatic triglyceride accumulation via endoplasmic reticulum stress-induced SREBP-1 activation is regulated by ceramide synthases. Exp Mol Med. 2019;51(11):1–16.

11. Chaurasia B, Tippetts TS, Mayoral Monibas R, Liu J, Li Y, Wang L, et al. Targeting a ceramide double bond improves insulin resistance and hepatic steatosis. Science. 2019;365(6451):386–92.

12. Raichur S, Wang ST, Chan PW, Li Y, Ching J, Chaurasia B, et al. CerS2 haploinsufficiency inhibits beta-oxidation and confers susceptibility to diet-induced steatohepatitis and insulin resistance. Cell metabolism. 2014;20(4):687–95.

13. Ichi I, Nakahara K, Fujii K, Iida C, Miyashita Y, Kojo S. Increase of ceramide in the liver and plasma after carbon tetrachloride intoxication in the rat. Journal of nutritional science and vitaminology. 2007;53(1):53–6.

14. Apostolopoulou M, Gordillo R, Koliaki C, Gancheva S, Jelenik T, De Filippo E, et al. Specific Hepatic Sphingolipids Relate to Insulin Resistance, Oxidative Stress, and Inflammation in Nonalcoholic Steatohepatitis. Diabetes Care. 2018;41(6):1235–43.

15. Luukkonen PK, Zhou Y, Sadevirta S, Leivonen M, Arola J, Oresic M, et al. Hepatic ceramides dissociate steatosis and insulin resistance in patients with non-alcoholic fatty liver disease. Journal of hepatology. 2016;64(5):1167–75.

16. Jung Y, Lee MK, Puri P, Koo BK, Joo SK, Jang SY, et al. Circulating lipidomic alterations in obese and non-obese subjects with non-alcoholic fatty liver disease. Aliment Pharmacol Ther. 2020;52(10):1603–14.

17. Carlier A, Phan F, Szpigel A, Hajduch E, Salem JE, Gautheron J, et al. Dihydroceramides in Triglyceride-Enriched VLDL Are Associated with Nonalcoholic Fatty Liver Disease Severity in Type 2 Diabetes. Cell Rep Med. 2020;1(9):100154.

18. Ooi GJ, Meikle PJ, Huynh K, Earnest A, Roberts SK, Kemp W, et al. Hepatic lipidomic remodeling in severe obesity manifests with steatosis and does not evolve with non-alcoholic steatohepatitis. Journal of hepatology. 2021.

19. Vvedenskaya O, Rose TD, Knittelfelder O, Palladini A, Wodke JAH, Schumann K, et al. Non-alcoholic fatty liver disease Stratification by Liver Lipidomics. Journal of lipid research. 2021:100104.

20. Satapathy SK, Kuwajima V, Nadelson J, Atiq O, Sanyal AJ. Drug-induced fatty liver disease: An overview of pathogenesis and management. Annals of hepatology. 2015;14(6):789–806.

21. Bedossa P, Poitou C, Veyrie N, Bouillot JL, Basdevant A, Paradis V, et al. Histopathological algorithm and scoring system for evaluation of liver lesions in morbidly obese patients. Hepatology (Baltimore, Md). 2012;56(5):1751–9.

22. Pfaffl MW. A new mathematical model for relative quantification in real-time RT-PCR. Nucleic Acids Res. 2001;29(9):e45.

23. Andrikopoulos S, Blair AR, Deluca N, Fam BC, Proietto J. Evaluating the glucose tolerance test in mice. American journal of physiology Endocrinology and metabolism. 2008;295(6):E1323–32.

24. Babiy B, Busto R, Pastor O. A normalized signal calibration with a long-term reference improves the robustness of RPLC-MRM/MS lipidomics in plasma. Analytical and bioanalytical chemistry. 2021;413(15):4077–90.

25. Folch J, Lees M, Sloane Stanley GH. A simple method for the isolation and purification of total lipides from animal tissues. The Journal of biological chemistry. 1957;226(1):497–509.

26. Younossi ZM, Koenig AB, Abdelatif D, Fazel Y, Henry L, Wymer M. Global epidemiology of nonalcoholic fatty liver disease-Meta-analytic assessment of prevalence, incidence, and outcomes. Hepatology (Baltimore, Md). 2016;64(1):73–84.

27. Loomba R, Friedman SL, Shulman GI. Mechanisms and disease consequences of nonalcoholic fatty liver disease. Cell. 2021;184(10):2537–64.

28. Farrell G, Schattenberg JM, Leclercq I, Yeh MM, Goldin R, Teoh N, et al. Mouse Models of Nonalcoholic Steatohepatitis: Toward Optimization of Their Relevance to Human Nonalcoholic Steatohepatitis. Hepatology (Baltimore, Md). 2019;69(5):2241–57.

29. Jahn D, Kircher S, Hermanns HM, Geier A. Animal models of NAFLD from a hepatologist’s point of view. Biochim Biophys Acta Mol Basis Dis. 2018.

30. Brown MS, Goldstein JL. Selective versus total insulin resistance: a pathogenic paradox. Cell metabolism. 2008;7(2):95–6.

31. Jensen T, Abdelmalek MF, Sullivan S, Nadeau KJ, Green M, Roncal C, et al. Fructose and sugar: A major mediator of non-alcoholic fatty liver disease. Journal of hepatology. 2018;68(5):1063–75.

32. Tomita K, Teratani T, Suzuki T, Shimizu M, Sato H, Narimatsu K, et al. Free cholesterol accumulation in hepatic stellate cells: mechanism of liver fibrosis aggravation in nonalcoholic steatohepatitis in mice. Hepatology (Baltimore, Md). 2014;59(1):154–69.

33. Tsuchida T, Lee YA, Fujiwara N, Ybanez M, Allen B, Martins S, et al. A simple diet- and chemical-induced murine NASH model with rapid progression of steatohepatitis, fibrosis and liver cancer. Journal of hepatology. 2018;69(2):385–95.

34. Gallou-Kabani C, Vige A, Gross MS, Rabes JP, Boileau C, Larue-Achagiotis C, et al. C57BL/6J and A/J mice fed a high-fat diet delineate components of metabolic syndrome. Obesity (Silver Spring). 2007;15(8):1996–2005.

35. Wong SK, Chin KY, Suhaimi FH, Fairus A, Ima-Nirwana S. Animal models of metabolic syndrome: a review. Nutrition & metabolism. 2016;13:65.

36. Podrini C, Cambridge EL, Lelliott CJ, Carragher DM, Estabel J, Gerdin AK, et al. High-fat feeding rapidly induces obesity and lipid derangements in C57BL/6N mice. Mamm Genome. 2013;24(5-6):240–51.

37. Zatloukal K, French SW, Stumptner C, Strnad P, Harada M, Toivola DM, et al. From Mallory to Mallory-Denk bodies: what, how and why? Exp Cell Res. 2007;313(10):2033–49.

38. Itoh M, Kato H, Suganami T, Konuma K, Marumoto Y, Terai S, et al. Hepatic crown-like structure: a unique histological feature in non-alcoholic steatohepatitis in mice and humans. PloS one. 2013;8(12):e82163.

39. Ioannou GN, Subramanian S, Chait A, Haigh WG, Yeh MM, Farrell GC, et al. Cholesterol crystallization within hepatocyte lipid droplets and its role in murine NASH. Journal of lipid research. 2017;58(6):1067–79.

40. Krishnan A, Abdullah TS, Mounajjed T, Hartono S, McConico A, White T, et al. A longitudinal study of whole body, tissue, and cellular physiology in a mouse model of fibrosing NASH with high fidelity to the human condition. American journal of physiology Gastrointestinal and liver physiology. 2017;312(6):G666–G80.

41. Wan J, Li J, Bandyopadhyay S, Kelly SL, Xiang Y, Zhang J, et al. Analysis of 1-Deoxysphingoid Bases and Their N-Acyl Metabolites and Exploration of Their Occurrence in Some Food Materials. Journal of agricultural and food chemistry. 2019;67(46):12953–61.

42. Sampath H, Miyazaki M, Dobrzyn A, Ntambi JM. Stearoyl-CoA desaturase-1 mediates the pro-lipogenic effects of dietary saturated fat. The Journal of biological chemistry. 2007;282(4):2483–93.

43. Senkal CE, Salama MF, Snider AJ, Allopenna JJ, Rana NA, Koller A, et al. Ceramide Is Metabolized to Acylceramide and Stored in Lipid Droplets. Cell metabolism. 2017;25(3):686–97.

44. van der Poorten D, Samer CF, Ramezani-Moghadam M, Coulter S, Kacevska M, Schrijnders D, et al. Hepatic fat loss in advanced nonalcoholic steatohepatitis: are alterations in serum adiponectin the cause? Hepatology (Baltimore, Md). 2013;57(6):2180–8.

45. van Koppen A, Verschuren L, van den Hoek AM, Verheij J, Morrison MC, Li K, et al. Uncovering a Predictive Molecular Signature for the Onset of NASH-Related Fibrosis in a Translational NASH Mouse Model. Cell Mol Gastroenterol Hepatol. 2018;5(1):83–98 e10.

46. Alsamman S, Christenson SA, Yu A, Ayad NME, Mooring MS, Segal JM, et al. Targeting acid ceramidase inhibits YAP/TAZ signaling to reduce fibrosis in mice. Science translational medicine. 2020;12(557).

47. Mauer AS, Hirsova P, Maiers JL, Shah VH, Malhi H. Inhibition of sphingosine 1-phosphate signaling ameliorates murine nonalcoholic steatohepatitis. American journal of physiology Gastrointestinal and liver physiology. 2017;312(3):G300–G13.

48. Kasumov T, Li L, Li M, Gulshan K, Kirwan JP, Liu X, et al. Ceramide as a mediator of non-alcoholic Fatty liver disease and associated atherosclerosis. PloS one. 2015;10(5):e0126910.

49. Raichur S, Brunner B, Bielohuby M, Hansen G, Pfenninger A, Wang B, et al. The role of C16:0 ceramide in the development of obesity and type 2 diabetes: CerS6 inhibition as a novel therapeutic approach. Molecular metabolism. 2019;21:36–50.

50. Hammerschmidt P, Ostkotte D, Nolte H, Gerl MJ, Jais A, Brunner HL, et al. CerS6-Derived Sphingolipids Interact with Mff and Promote Mitochondrial Fragmentation in Obesity. Cell. 2019;177(6):1536–52 e23.

51. Nicholson RJ, Poss AM, Maschek JA, Cox JE, Hopkins PN, Hunt SC, et al. Characterizing a Common CERS2 Polymorphism in a Mouse Model of Metabolic Disease and in Subjects from the Utah CAD Study. J Clin Endocrinol Metab. 2021;106(8):e3098–e109.

52. Alonso A, Goni FM. The Physical Properties of Ceramides in Membranes. Annu Rev Biophys. 2018;47:633–54.

53. Wigger L, Cruciani-Guglielmacci C, Nicolas A, Denom J, Fernandez N, Fumeron F, et al. Plasma Dihydroceramides Are Diabetes Susceptibility Biomarker Candidates in Mice and Humans. Cell reports. 2017;18(9):2269–79.

54. Mayo R, Crespo J, Martinez-Arranz I, Banales JM, Arias M, Minchole I, et al. Metabolomic-based noninvasive serum test to diagnose nonalcoholic steatohepatitis: Results from discovery and validation cohorts. Hepatology communications. 2018;2(7):807–20.

55. Liebisch G, Ekroos K, Hermansson M, Ejsing CS. Reporting of lipidomics data should be standardized. Biochim Biophys Acta Mol Cell Biol Lipids. 2017;1862(8):747–51.

56. Castro RE, Diehl AM. Towards a definite mouse model of NAFLD. Journal of hepatology. 2018;69(2):272–4.

57. Anand AC. Nutrition and Muscle in Cirrhosis. J Clin Exp Hepatol. 2017;7(4):340–57.

58. Masoodi M, Gastaldelli A, Hyotylainen T, Arretxe E, Alonso C, Gaggini M, et al. Metabolomics and lipidomics in NAFLD: biomarkers and non-invasive diagnostic tests. Nat Rev Gastroenterol Hepatol. 2021;18(12):835–56.

