## Supplementary_Figure_Legends for "Accumulation of dihydrosphingolipids and neutral lipids is related to steatosis and fibrosis damage in human and animal models of non-alcoholic fatty liver disease"

**Suppl. Fig. 1. Overview of sphingolipid metabolism in NAFLD.** Ceramides (Cer) and dihydroceramides (dhCer) are derived from condensation of serine and palmitic acid which is also precursor other storage neutral lipids (TG and CE). In the pre-diabetic insulin resistant conditions, the oversupply of FFA stimulates the synthesis of neutral lipids in peripheral tissues. In early steatosis (NAFL) the liver is able to accommodate the increase of FFA in the form of neutral lipids, but at the same time, triggers the synthesis of dhCer and Cer. Other (dihydro)sphingolipid species and acylceramide (ACer) species could also be synthesized by condensation of different substituents (Z) whose influence on NAFLD progression is unknown. FFA, free fatty acids; TG, triglyceride; CE, Cholesteryl ester; ACer, 1-o-Acylceramide; Cer, ceramide; HexCer, hexosylceramide; SM, Sphingomyelin; dhCer, dihydroceramide; dhHexCer, dihydrohexosylceramide dhSM, dihydrosphingomyelin; Z, phosphorylcholine and hexoses (glucose and galactose).

**Suppl. Fig. 2. Experimental design used in the mice model of NAFLD.**

**Suppl. Fig. 3. Hepatic stellate cell activation in the mice model.** Immunostaining for α-SMA (ACTA2) in liver sections from representative mice treated with Original magnification x100. Representative liver from mice on normal chow diet for 22 weeks (CNT 22w); high-fat diet for 22 weeks (HFD 22w); HFD for 30 weeks (HFD 30w); HFD for 40 weeks (HFD 40w), HFD 22w+$CCl_4$ for 6 weeks ($CCl_4$ 6w) and HFD 22w+$CCl_4$ for 10 weeks ($CCl_4$ 10w). Original magnification x100. Black arrows marked the appearance of hepatic crown-like structures (hCLS). α-SMA, alpha-smooth muscle actin; $CCl_4$, carbon tetrachloride; HFD, high-fat diet.

**Suppl. Fig. 4. Classification of mice following the SAF score.** (A) The histological scores were obtained from liver sections stained with hematoxylin & eosin (steatosis, ballooning and inflammation) and sirius red (fibrosis). (B) Mice were classified according to the SAF algorithm into: non-NAFLD, simple steatosis (NAFL), steatohepatitis without significant fibrosis, stage of fibrosis F0-F1 (NASH) and steatohepatitis + stage of fibrosis F2-F4 (NASH-fibrosis). Values inside bars represent the grading/staging for the score and the number of animals falling on each category. S, steatosis; A, activity; F, fibrosis. n, number of animals;

**Suppl. Fig. 5. Relationship between the fibrosis staging and steatosis grading with liver lipid changes.** Quantitative analysis of lipid species from 10 lipid clases, were determined in livers of mice grouped by the fibrosis stage. The values of $Log_2$-fold change of each animal (columns) and each lipid specie (rows) is plotted in the in the heatmap and ordered by the hierarchical clustering of individuals according to the steatosis score (columns). TG, triglyceride; CE, Cholesteryl ester; FC, free cholesterol; ACer, 1-o-Acylceramide; Cer, ceramide; HexCer, hexosylceramide; SM, Sphingomyelin; dhCer, dihydroceramide; dhHexCer, dihydrohexosylceramide dhSM, dihydrosphingomyelin.

**Suppl. Fig. 6. Gene expression of *"de novo"* sphingolipid synthesis enzymes in liver of NAFLD mice.** Gene expression levels of *Sptlc1-3, Cers2, Cers4-6, Degs1, Sgms1-2* in the liver of mice were classified according to the SAF algorithm into: non-NAFLD, simple steatosis (NAFL), steatohepatitis without significant fibrosis, stage of fibrosis F0-F1 (NASH) and steatohepatitis + stage of fibrosis F2-F4 (NASH-fibrosis) Data represent the mean ± SD of n=5-10 samples per group. *p <0.05, **p <0.01, ***p <0.001. ****p <0.0001.

**Suppl. Fig. 7. Classification of NAFLD patients following the SAF score.** The histological scores were obtained from liver sections stained with hematoxylin & eosin (steatosis, ballooning and inflammation) and Masson’s trichrome (fibrosis). Liver from the (A) Bariatric surgery patients and (B) Full patient cohort for plasma lipidomics, were classified according to the SAF algorithm into: non-NAFLD, simple steatosis (NAFL), steatohepatitis without significant fibrosis, stage of fibrosis F0-F1 (NASH) and steatohepatitis + stage of fibrosis F2-F4 (NASH-fibrosis). Values inside bars represent the grading/staging for the score and the number of animals falling on each category. S, steatosis; A, activity; F, fibrosis. n, number of patients.

**Suppl. Fig. 8. Correlation of triglyceride species with ceramides and dihydroceramides in the liver of NAFLD patients.** (A) Correlation plot between representative species of Cer and TG species. (B) Correlation plot between representative species of dhCer and TG species. *p <0.05, *p <0.01, ***p <0.001 and ****p <0.001. R, Pearson’s correlation coefficient; ns, non-significant; TG, triglyceride; Cer, ceramide; dhCer, dihydroceramide; NASF; NASH-fibrosis.

**Suppl. Fig. 9. Quality control assurance of lipidomic analyses. (**A) Principal components analysis (PCA) scores plot of the quality controls (QCs) included on every batch. Same color corresponds to QC processed on the same batch of analysis. Between batch and within batch Relative Standard Deviation (RSD) obtained in the analysis of lipid classes in the QC-low (B), QC-high (C) and QC-medium (D).
