## Supplementary_Tables for "Accumulation of dihydrosphingolipids and neutral lipids is related to steatosis and fibrosis damage in human and animal models of non-alcoholic fatty liver disease"

**Suppl. Table S1. Macroscopic and biochemical characteristics of mice classified by the SAF score.** Mice were classified according to their liver histological findings, and with independence of their time on diet or administration of CCl4, into four groups: non-NAFLD, simple steatosis (NAFL), steatohepatitis without significant fibrosis, stage of fibrosis F0-F1 (NASH) and steatohepatitis + stage of fibrosis F2-F4 (NASH-fibrosis). Values are expressed in mean ± SD (N, number of animals). (a) Significant, $p<0.05$ from non-NAFLD. (b) Significant, $p<0.05$ from NAFL. (c) Significant, $p<0.05$ from NASH. (d) Significant, $p<0.05$ from NASH-fibrosis. ALT, alanine aminotransferase; AST, aspartate aminotransferase; LW, Liver weight, BW, body weight; HDLc, high-density lipoprotein cholesterol; FFA, free fatty acids.

| **Variable** | **non-NAFLD** | **NAFL** | **NASH** | **NASH-fibrosis** |
|---|---|---|---|---|
| Weight, g | 33.7 ± 0.5 (7)$^{b,c,d}$ | 38.6 ± 1.1 (17)$^{a}$ | 44.0 ± 2.7 (9)$^{a}$ | 39.0 ± 1.8 (24)$^{a}$ |
| Liver weight, g | 1.40 ± 0.04 (7)$^{b,c,d}$ | 1.93 ± 0.11 (17)$^{a,c}$ | 2.85 ± 0.36 (9)$^{a,b}$ | 2.38 ± 0.37 (24)$^{a}$ |
| LW/BW, % | 4.03 ± 0.21 (7)$^{b,c,d}$ | 4.94 ± 0.20 (17)$^{a}$ | 6.28 ± 0.53 (9)$^{a,b}$ | 5.53 ± 0.62 (24)$^{a}$ |
| Glucose, mg/dL | 149.9 ± 8.6 (7)$^{b,c}$ | 196.1 ± 8.7 (17)$^{a,c,d}$ | 232.2 ± 11.4 (9)$^{a,b,d}$ | 165.6 ± 6.6 (24)$^{b,c}$ |
| ALT, IU/mL | 17.1 ± 1.1 (7)$^{b,c,d}$ | 87.5 ± 24.3 (17)$^{a}$ | 108.8 ± 24.0 (9)$^{a}$ | 151.2 ± 31.7 (24) |
| AST, IU/mL | 83.86 ± 9.32 (7)$^{c,d}$ | 117.59 ± 17.56 (17)$^{c}$ | 137.75 ± 14.83 (9)$^{a}$ | 236.67 ± 34.22 (24)$^{a,b,c}$ |
| Triglycerides, mg/dL | 67.3 ± 9.4 (7)$^{b,d}$ | 37.41 ± 2.09 (17)$^{a,d}$ | 47.75 ± 6.15 (9)$^{d}$ | 31.12 ± 1.55 (24)$^{a,b,c}$ |
| Cholesterol, mg/dL | 88.7 ± 9.7 (7)$^{b,c,d}$ | 131.8 ± 6.8 (17)$^{a}$ | 150.4 ± 12.3 (9)$^{a}$ | 129.3 ± 15.8 (24)$^{a}$ |
| HDLc, mg/dL | 53.4 ± 5.79 (7)$^{b,c}$ | 72.1 ± 3.6 (16)$^{a,d}$ | 78.5 ± 5.3 (9)$^{a,d}$ | 57.4 ± 4.9 (24)$^{b,c}$ |
| Protein, g/dL | 3.46 ± 0.30 (7)$^{b,c,d}$ | 4.13 ± 0.07 (15)$^{a,d}$ | 4.61 ± 0.13 (9)$^{a,d}$ | 4.19 ± 0.11 (24)$^{a,b,c}$ |
| Urea, mg/dL | 25.7 ± 2.0 (7)$^{b,c,d}$ | 38.0 ± 1.6 (17)$^{a}$ | 33.6 ± 2.7 (9)$^{a}$ | 36.8 ± 1.0 (24)$^{a}$ |
| FFA, nmol/mL | 0.42 ± 0.02 (5)$^{b,c}$ | 0.54 ± 0.04 (7)$^{a,d}$ | 0.57 ± 0.03 (6)a,d | 0.41 ± 0.02 (10)$^{b,c}$ |
| ***Liver lipid classes*** | | | | |
| TG, nmol/mg | 49.7 ± 8.6 (7)$^{b,c,d}$ | 94.7 ± 4.7 (17)$^{a}$ | 95.1 ± 8.9 (9)$^{a}$ | 98.4 ± 8.5 (24)$^{a}$ |
| CE, nmol/mg | 7.7 ± 0.5 (7)$^{b,c,d}$ | 224.5 ± 24.6 (17)$^{a}$ | 241.7 ± 36.8 (9)$^{a}$ | 167.8 ± 18.4 (24)$^{a,b}$ |
| FC, nmol/mg | 38.3 ± 2.2 (7)$^{b,c,d}$ | 71.9 ± 3.1 (17)$^{a,d}$ | 79.1 ± 7.5 (9)$^{a}$ | 61.5 ± 3.1 (24)$^{a,b,c}$ |
| ACer, nmol/mg | 0.005 ± 0.001 (7) | 0.043 ± 0.005 (17) | 0.05 ± 0.009 (9) | 0.04 ± 0.006 (24) |
| Cer, nmol/mg | 1.21 ± 0.11 (7)$^{c}$ | 1.30 ± 0.08 (17)$^{c,d}$ | 1.95 ± 0.28 (9)$^{a,b}$ | 1.24 ± 0.12 (24)$^{c}$ |
| HexCer, nmol/mg | 0.29 ± 0.03 (7)$^{b,c,d}$ | 0.67 ± 0.05 (17)$^{a}$ | 0.73 ± 0.06 (9)$^{a}$ | 1.04 ± 0.10 (24)$^{a}$ |
| SM, nmol/mg | 8.56 ± 0.41 (7)$^{d}$ | 7.79 ± 0.82 (17)$^{d}$ | 7.98 ± 1.41 (9)$^{d}$ | 13.01 ± 1.50 (24)$^{a,b,c}$ |
| dhCer, nmol/mg | 0.024 ± 0.003 (7)$^{a,b,c}$ | 0.039 ± 0.003 (17)$^{a,d}$ | 0.046 ± 0.006 (9)$^{a}$ | 0.049 ± 0.005 (24)$^{a,b}$ |
| dhHexCer, nmol/mg | 0.005 ± 0.001 (7)$^{a,b,c}$ | 0.008 ± 0.001 (17)$^{a,d}$ | 0.010 ± 0.001 (9)$^{a}$ | 0.012 ± 0.001 (24)$^{a,b}$ |
| dhSM, nmol/mg | 0.36 ± 0.02 (7)$^{a,b,c}$ | 0.78± 0.04 (17)$^{a,d}$ | 0.79 ± 0.08 (9)$^{a,d}$ | 1.10 ± 0.11 (24)$^{a,b,c}$ |
| LPC, nmol/mg | 3.33 ± 0.43 (7)$^{c}$ | 2.71 ± 0.22 (17) | 2.16 ± 0.29 (9)$^{a,d}$ | 2.99 ± 0.26 (24)$^{c}$ |
| PC, nmol/mg | 88.1 ± 3.8 (7)$^{b,d}$ | 74.4 ± 5.3 (17)$^{a}$ | 74.4 ± 7.9 (9) | 64.8 ± 7.9 (24)$^{a}$ |
| PE, nmol/mg | 62.1 ± 3.0 (7)$^{b,c,d}$ | 43.9 ± 2.3(17)$^{a}$ | 44.6 ± 3.8 (9)$^{a}$ | 50.5 ± 4.7 (24)$^{a}$ |
| FFA, nmol/mg | 0.16 ± 0.01 (5)$^{c,d}$ | 0.22 ± 0.05 (7)$^{c,d}$ | 0.49 ± 0.05 (6)$^{a,b}$ | 0.44 ± 0.03 (10)$^{a,b}$ |
