## Supplementary_Methods for "Accumulation of dihydrosphingolipids and neutral lipids is related to steatosis and fibrosis damage in human and animal models of non-alcoholic fatty liver disease"

(1) Servicio de Bioquímica Clínica, UCA-CCM, HU Ramón y Cajal-IRYCIS, Madrid, Spain.
(2) Instituto Murciano de Investigación Biosanitaria (IMIB-Arrixaca), Murcia, Spain.
(3) Servicio de Bioquímica-Investigación, HU Ramón y Cajal-IRYCIS, Madrid, Spain.
(4) Servicio de Gastroenterología, HU Ramón y Cajal-IRYCIS, Madrid, Spain.
(5) Servicio de Cirugía General. HCU Virgen de la Victoria. Málaga, Spain.
(6) CIBER de Fisiología de la Obesidad y Nutrición (CIBEROBN), ISCIII, Spain.
(7) Servicio de Anatomía Patológica. HU Ramón y Cajal-IRYCIS, Madrid, Spain.
(8) Departamento de Cirugía General y Aparato Digestivo, HU Virgen de la Arraixaca, Murcia, Spain.
(9) CIBER de Enfermedades Hepáticas y Digestivas (CIBEREHD), ISCIII, Spain.

*Animal housing conditions*

Mice were housed in the animal facility of Hospital Universitario Ramón y Cajal (Num. Reg: ES 280790000092) in a temperature-controlled room on a 12 light/dark cycle with *ad libitum* access to food and water. All interventions were performed during the light cycle and in accordance with legislation in Spain (RD 53/2013) and the European Directive (2010/63/EU) and approved by the Ethics Committee of the Ramón y Cajal Hospital, Madrid (ES-280790002001).

*Diets, treatments and sacrificing conditions*

Six week old C57BL/6J male mice (Charles River) received either standard rodent chow diet (SAFE A04; SAFE, Rosenberg, Germany; with 19.3 kcal% protein, 72.4 kcal% carbohydrate, and 8.4 kcal% fat) or a high-fat diet (HFD) (D12108C; Research Diets, Inc, New Brunswick, 205 NJ; with 20 kcal% protein, 40 kcal% carbohydrate, and 40 kcal% fat, supplemented with 1.25% Cholesterol by weight) and a high sugar solution (23.1 g/L D-fructose (Sigma-Aldrich, F0127) and 18.9 g/L D-glucose (Sigma-Aldrich, G8270) in drinking water up to 40 weeks of age (32 weeks on HFD diet). At 22 weeks a subset group of animals received a weekly intraperitoneal injection of $CCl_4$ (Sigma-Aldrich, 289116-100ML) at final dose of 0.2 $\mu$l (0.32 $\mu$g of $CCl_4$)/gr of body weight, diluted (1:10) in mineral oil (Sigma-Aldrich, M5310-500ML) for 6 and 10 weeks, to induce a NASH-fibrosis phenotype (1). Animals were sacrificed at predetermined time points (**Suppl. Fig 2)**, food was removed 6h before death. Mice were anesthetized with sevoflorane (2% in oxygen flux at 1L/min). Blood was drawn by cardiac puncture and plasma obtained after centrifugation for biochemistry assays. The liver was extracted and weighed. The median lobe was preserved in 10% buffered formalin for histology and immunohistochemistry. Additional 2-3mm cubes of tissue were collected into plastic cryotubes, and immediately frozen in nitrogen to perform lipidomics, western-blot protein and gene expression analyses.

*Mouse cheek-nick blood collection*

Plasma samples were taken immediately before putting animals on diet at basal (6 weeks) and at 4, 8, and 16 weeks of age. Blood was withdrawn from the submandibular vein (after 12h fasting) performing a nick on the cheek of animals. Five drops (~100µL) were collected in a tube with 10µL containing a solution of EDTA-$K_3$ salt (4.5 gr/100mL). Plasma was isolated after centrifugation and separated and freezed at for biochemistry assays.

*Histology of patient liver biopsies*

Intraoperative wedge liver biopsies from bariatric surgery patients, at least one centimeter in depth, were taken. One section was snap frozen and stored at -80C for lipidomic analysis, and one section was formalin fixed paraffin-embedded for histological assessment. Percutaneous or transjugular liver biopsies specimens (1 mm diameter and 1.5 mm length) were taken from the non-bariatric surgery patients included in the plasma lipidomic cohort. The minimal set of staining included hematoxylin and eosin (H&E), Masson trichrome, Periodic acid–Schiff (PAS), Perls and reticulin staining. All biopsies were reviewed by trained liver pathologist from the recruitment centers.

The grade of steatosis (S, from S0 to S3 according to the previously defined thresholds of steatosis amount), the grade of activity (A from A0 to A4 by addition of grades of ballooning and lobular inflammation, each graded from 0 to 2 as defined before) and the stage of fibrosis (F from F0 to F4), the latter according to NASH-CRN staging system(2), were summarized according to the SAF (steatosis, activity and fibrosis) classification system proposed by Bedossa (3). Classification of patients into non-NAFLD (healthy obese), NAFL (simple steatosis), and non-alcoholic steatohepatitis (NASH), was done following the algorithm proposed in the same work. Subclassification of NASH patients into NASH without and with significant fibrosis (NASH-fibrosis) was done according to a fibrosis stage F ≥ 2. Patients with S=0 & A ≥ 1 (*burn-out* NASH) were classified as NASH or NASH-Fibrosis following the same criteria.

*Histology of mouse liver biopsies*

Mice livers were formalin-fixed and embedded on paraffin. Liver sections were stained with haematoxylin and eosin, and picrosirius red (Direct Red 80, Sigma-Aldrich), and evaluated by an experienced pathologist (P.C). The scoring was obtaining adapting and scored adapting the SAF (Steatosis, Activity and Fibrosis) classification proposed by Bedossa (3). The histological scoring was performed in blind.

*Immunohistochemistry*

Liver sections (5 µum) were deparaffinized and rehydrated, and antigen retrieval was achieved by heat treatment in a pressure cooker for 2 min in 10 mM citrate buffer (pH 6.5). Next, endogenous peroxidase was blocked, and the sections were incubated either with anti-SMA (1:500) (ACTA2 / $\alpha$-SMA) (Biorbyt ltd, ref: orb155585) overnight at room temperature. the sections were developed using Master Polymer Plus Detection System (Peroxidase) (Master Diagnostica) according to the manufacturer’s instructions. The sections were counterstained with hematoxylin.

*Western-blot protein expression*

Liver tissue (10-30 mg) was added to a 1.5 mL screwcap plastic tubes containing 800 µL of lysis buffer solution: Tissue homogenization were performed in a solution of 50 mM Tris–HCl pH 7.5, 125 mM NaCl, 1% Nonident-P40, 5 mM NaF, 1.4 mM $Na_4O_7P_2$, 1 mM $Na_3VO_4$, and protease inhibitor (Pierce Biotechnology, Inc., Rockford, IL, USA), using a BeadBug homogenizer (Benchmark Scientific #D1036*) at 3000rpm with 1.5 mm Triple-

Pure High Impact Zirconium Beads (Benchmark Scientific #D1032-15). Afterwards the tissue extracts were diluted to 1mg/ml to measure protein concentration by a bicinchoninic acid assay (BCA, Pierce-Thermo Fisher Scientific #23227). Protein expression was measured loading 40 µg of protein onto 10% PAGE-SDS gels for electrophoresis at a constant voltage of 100 V at 4°C. Next, the transference of proteins was performed at 100 V for 1 hour at 4°C onto nitrocellulose membranes (Merck-Millipore Corp., Darmstadt, Germany). After blocking, the blots were incubated with one of the following specific antibodies: HSP90 (BD Biosciences, ref: 610418); ACTA2/$\alpha$-SMA (Biorbyt ltd, ref: orb155585); PLIN2 (Novus, ref: NB110-40877); p62 (Abcam, ref: Ab56416); Col1A1 (Cell Signalling, ref: 84336). Subsequently, a secondary antibody conjugated to IRDye 800 CW or IRDye 680 LT (LI-COR, Lincoln, NE, USA) were used. The membranes were analyzed using the Odyssey Infrared Imaging System (LI-COR Bioscience). Densitometric analysis was performed using the Odyssey Infrared Imaging System V.3.0

*Real time PCR gene expression*

Total RNA from liver tissue (5-20mg) was extracted with RNeasy Plus Universal Mini Kit using the QIAcube automated purification system (QIAGEN), according to the manufacturer's recommendation. The liver samples (5-20mg) were homogenized using 1.6 mm steel bead (Cultek #F7SSB16-RNA) on BeadBug homogenizer (Benchmark Scientific #D1036). RNA quality and quantity were assessed using the NanoDrop ND-1000 (NanoDrop Technologies, Wilmington, DE) and agarose gel electrophoresis. 500 ng of total RNA were reverse transcribed with random hexamers using the PrimeScript RT reagent kit (Takara). Real-time PCR amplification was performed on a LightCycler 480 II using the SYBR Green I Master kit (Roche) on 384 well-plates. All determinations were triple analyzed, and the thermocycler protocol consisted of a preincubation step at 95°C for 5 min, followed by 45 cycles at 95°C for 10 s, 60°C for 10 s, and 72°C for 10 s. The melting curves were evaluated for each sample to ensure the presence of a single product. For each probe, the efficiency of the amplification reaction was tested by serial dilutions of cDNA, ensuring that the amplification efficiency was linear and was close to 2. The PCR reaction products were also separated on a 2% agarose gel to confirm the amplicon size. The results were analyzed using the relative quantification method described by Pfaffl (4), which takes into account the efficiency of the reaction for the target gene and the invariable control. Three housekeeping genes (*Rn18s, Cypb, and Rplp0*) were used as invariant endogenous controls for each sample. For comparison and statistical analysis, each data obtained was normalized to the expression of the control group. The sequence of primers used in this study can be found in **Suppl. Table S2**.

*Oral glucose tolerance test*

Oral glucose tolerance test (OGTT) in mice were performed after 5h fasting(5). An oral solution of glucose at 0.5g/ml were administered on mice using a gavage needle at 2g glucose/kg mouse weight. Blood glucose measurements were done at 0, 15, 30, 60, and 120 min, performing a nick in the tail of animals, with a glucometer (Performa-Accu-Chek, Roche).

*Blood biochemistry assays*

Glucose (Glu), Aspartate aminotransferase (AST), Alanine aminotransferase (ALT), total cholesterol (Chol), Total total triacylglycerols (Trig), HDL cholesterol (HDLc), Insulin, and other blood clinical chemistry measurements were measured in an Architect ci16000 system (Abbott diagnostics). HbA1c was done on a Hb9210 Premier (A.Menarini diagnostics). Free Fatty Acid (FFA) in plasma and liver tissue measurements were done by and ELISA kit (Elabscience). The PNIIIP, HA and TIMP1 in plasma were analysed on an ADVIA-Centaur XPT system (Siemens healthineers) and CK18 by ELISA kit (PEVIVA).

*Lipidomics of liver tissue and plasma*

Liver tissue (10-30 mg) was added to a 1.5 mL screwcap plastic tubes containing 800 µL of lysis buffer solution: 50 mM Tris–HCl pH 7.5, 125 mM NaCl, 5 mM NaF,1.4 mM $Na_4O_7P_2$, 1 mM $Na_3VO_4$, and protease inhibitor (Pierce Biotechnology, Inc., Rockford, IL, USA), and 1.5 mm zirconium beads (Benchmark Scientific #D1032-15). The tissues were disgregated on a BeadBug-6 Homogenizer (Benchmark D1036-E, Bechmark Scientific) at 4500 rpm, in 3 cycles of 90 s. The homogenate was further sonicated 40s at 10% of amplitude and centrifuged at 600 × g for 1 min at 4°C. An aliquot of the resultant supernatant was diluted 1:10 for total protein determination by the bicinchoninic acid (BCA) protein assay (Pierce Biotechnology Inc.). The rest of the protein extract was stored at -80ºC until lipidomic analysis.

The lipidomic analysis was done following a previously validated methodology (6). Briefly, lipids from liver (equivalent to 250 µg of quantified total protein) or plasma (50 µL) were extracted following the Folch's method (7). Ten microliters of an internal standard mix **(**IS-mix**)** was added before the lipid extraction to all samples obtain the relative molar quantification. The concentrations of the IS-mix are given in **Suppl. Table S3.** The Lipid extracts were dried and suspended in 250 µL of acetonitrile/isopropanol (1:1, vol:vol), sonicated for 10 min, and then transferred to an injection vial. Five microliters of the lipid extract were injected on an LC system Eksigent UltraLC-100 (AB-Sciex LLP, Framingham, MA, USA). The species were separated on a Kinetex C18 column (100 × 2.1 mm, 1.7 µm; Phenomenex, Macclesfield, UK) maintained at 55ºC. Elution was carried out using a system consisting of solvent A (60% acetonitrile in water, 10 mM ammonium formate) and solvent B (90% isopropyl alcohol in acetonitrile, 10 mM ammonium formate) and a linear gradient from 60% A to 100% B in 12 min and 100% B to 60% A in 8 min at a flow rate of 0.4 mL/min. The detection of lipid species was carried out following a targeted approach setting MRM transitions for each lipid species at their retention times (**Suppl. Table S4)**.

The detection of lipid classes was done on a QTrap 4000 (AB-Sciex LLP, Framingham, MA, USA) with Analyst 1.6.2 software. Nitrogen was employed as the drying gas at a temperature of 500ºC, the curtain gas was set at 30 psi, and the ion source gas was set at 50 psi. CE and FC were analysed using the atmospheric pressure chemical ionization (APCI) source in the positive ion mode. The LC-MS/MS peak chromatograms were processed using Skyline software version 4.1 (8, 9). The lipid species were quantified according to a matrix-matched single-point calibration with internal standardization schema as previously described (6). The level of absolute quantification achieved by our methodology is level 3 following the Lipidomics Standard Initiative guidelines (10).

*Quality control assurance of lipidomic analysis*

Quality control assurance of lipidomic analysis was performed on regular basis for every lipidomic analysis using a calibration-based methodology described in (6). In summary, we used the following parameters to evaluate the reproducibility of quality control samples lipid concentration through time: Relative Standard Deviation (RSD) applied to lipid species and class sum within or between batches of analysis; Principal Component Analysis (PCA) for global evaluation of the quality control samples (**Suppl. Fig 9**).

**Suppl. Table S2. List of specific primers for mRNA amplification in liver mice.**

| Gene | Forward primer (5'-3') | Reverse primer (5'-3') |
|---|---|---|
| *Acta2* | GACTCTCTTCCAGCCATCTTTC | GACAGGACGTTGTTAGCATAGA |
| *Cd36* | GATGACGTGGCAAAGAACAG' | TCCTCGGGGTCCTGAGTTAT |
| *Cers2* | AGAGTGGGCTCTCTGGACG | CCAGGGTTTATCCACAGTGAC |
| *Cers4* | GACCGTGATGGCCTGGTGTT | TCTCCTGATTGGATCCTGCA |
| *Cers5* | GACTGCTTCCAAAGCCTTGAG | GCAGTTGGCACCATTGCTAG |
| *Cers6* | AAGCCAATGGACCACAAACT' | TGCTTGGAGAGCCCTTCTAAT |
| *Cypb* | TGGAGAGCACCAAGACAGACA | TGCCGGAGTCGACAATGAT |
| *Degs1* | AGCCACCTTGCTTGGCCTGG | TGCGATCTTCCTCACCATGGG |
| *Mcp1* | TCTGGGCCTGCTGTTCACA | GGATCATCTTGCTGGTGAATGA |
| *Rplp0* | AAGCGCGTCCTGGCATTGTCT | CCGCAGGGGCAGCAGTGGT |
| *Scd1* | CTGTACGGGATCATACTGGTTC | GCCGTGCCTTGTAAGTTCTG |
| *Sgms1* | TGGGCGTTTTCTATTTGCGAA | GTACAGCGTGCCAACTATGC |
| *Sgms2* | GACTTCCTCTTCAGCGGCC | TGCGCTACGAGAATGCAGAT |
| *Sptlc1* | ACGAGGCTCCAGCATACCAT | TCAGAACGCTCCTGCAACTTG |
| *Sptlc2* | AACGGGGAAGTGAGGAACG | CAGCATGGGTGTTTCTTCAAAAG |
| *Tgfb1* | TGGAGCAACATGTGGAACTC | GTCAGCAGCCGGTTACCA |
| *Tnfa* | GCCTCTTCTCATTCCTGCTTG | CTGATGAGAGGGAGGCCATT |
| *Rn18s* | CCAGTAAGTGCGGGTCATAAGC | CCTCACTAAACCATCCAATCGG |

**Suppl. Table S3. Concentration of lipid species in the IS-mix used in lipidomic experiments**

| Standard | Vendor & Reference | Conc. IS-mix (nmol/mL) |
|---|---|---|
| ACer 35:1 | Avanti Lipids, #860526 | 0.5 |
| CE 16:0-d7 | Avanti Lipids, #700149 | 200 |
| Cer 42:1-d7 | Avanti Lipids, #860678 | 5 |
| FC-d7 | Avanti Lipids, #700041 | 200 |
| HexCer 30:1 | Avanti Lipids, #860543 | 100 |
| LPC 17:0 | Avanti Lipids, #855676 | 100 |
| PC 28:2 | Avanti Lipids, #850347 | 200 |
| PE 32:2 | Avanti Lipids, #850706 | 100 |
| SM 30:1 | Avanti Lipids, #860583 | 100 |
| dhSM 30:0 | Avanti Lipids, #860683 | 20 |
| TG 46:2 | Larodan, #34-1018-7 | 50 |
| dhCer 35:0 | Cayman Chemical, #24427 | 2 |

**Suppl. Table S4. List of MRM transitions, retention time and mass spectrometer settings.**

| Q1 Mass (Da) | Q3 Mass (Da) | RT (min) | Sum specie | Source (mode) | DP (volts) | EP (volts) | CE (volts) | CXP (volts) |
|---|---|---|---|---|---|---|---|---|
| 816.8 | 264.3 | 12 | IS-ACer 35:1 | ESI (+) | 115 | 10 | 55 | 14 |
| 802.8 | 264.3 | 11.8 | ACer 34:1 | ESI (+) | 115 | 10 | 55 | 14 |
| 804.801 | 264.3 | 12.2 | ACer 34:0 | ESI (+) | 115 | 10 | 55 | 14 |
| 830.8 | 264.3 | 12.3 | ACer 36:1 | ESI (+) | 115 | 10 | 55 | 14 |
| 914.8 | 264.3 | 13.6 | ACer 42:1 | ESI (+) | 115 | 10 | 55 | 14 |
| 912.801 | 264.3 | 13.2 | ACer 42:2 | ESI (+) | 115 | 10 | 55 | 14 |
| 538.6 | 264 | 6.3 | Cer 34:1 | ESI (+) | 66 | 10 | 57 | 10 |
| 566.6 | 264 | 7.3 | Cer 36:1 | ESI (+) | 66 | 10 | 57 | 10 |
| 594.8 | 264 | 8.3 | Cer 38:1 | ESI (+) | 66 | 10 | 57 | 10 |
| 622.8 | 264 | 9.2 | Cer 40:1 | ESI (+) | 66 | 10 | 57 | 10 |
| 650.8 | 264 | 10 | Cer 42:1 | ESI (+) | 66 | 10 | 57 | 10 |
| 648.8 | 264 | 9.2 | Cer 42:2 | ESI (+) | 66 | 10 | 57 | 10 |
| 540.5 | 266 | 6.7 | dhCer 34:0 | ESI (+) | 66 | 10 | 57 | 10 |
| 568.5 | 266 | 7.7 | dhCer 36:0 | ESI (+) | 66 | 10 | 57 | 10 |
| 596.2 | 266 | 8.7 | dhCer 38:0 | ESI (+) | 66 | 10 | 57 | 10 |
| 624.5 | 266 | 9.5 | dhCer 40:0 | ESI (+) | 66 | 10 | 57 | 10 |
| 652.6 | 266 | 10 | dhCer 42:0 | ESI (+) | 66 | 10 | 57 | 10 |
| 650.6 | 266 | 9.5 | dhCer 42:1 | ESI (+) | 66 | 10 | 57 | 10 |
| 700.8 | 264 | 5.1 | HexCer 34:1 | ESI (+) | 66 | 10 | 57 | 10 |
| 728.6 | 264 | 6.1 | HexCer 36:1 | ESI (+) | 66 | 10 | 57 | 10 |
| 756.8 | 264 | 7.1 | HexCer 38:1 | ESI (+) | 66 | 10 | 57 | 10 |
| 784.8 | 264 | 8.1 | HexCer 40:1 | ESI (+) | 66 | 10 | 57 | 10 |
| 812.8 | 264 | 9 | HexCer 42:1 | ESI (+) | 66 | 10 | 57 | 10 |
| 657.8 | 271 | 9.9 | IS-Cer 42:1 (d7) | ESI (+) | 125 | 10 | 53 | 10 |
| 554.5 | 266.2 | 7.2 | IS-dhCer 35:0 | ESI (+) | 66 | 10 | 57 | 10 |
| 649.5 | 184 | 3 | IS-dhSM 30:0 | ESI (+) | 106 | 10 | 35 | 15 |
| 644.5 | 264 | 3.1 | IS-HexCer 30:1 | ESI (+) | 66 | 10 | 57 | 10 |
| 510.4 | 184 | 1 | IS-LPC 17:0 | ESI (+) | 90 | 10 | 40 | 15 |
| 674.6 | 184 | 2.4 | IS-PC 28:2 | ESI (+) | 90 | 10 | 40 | 15 |
| 688.6 | 547.6 | 4.2 | IS-PE 32:2 | ESI (+) | 110 | 10 | 35 | 15 |
| 647.6 | 184 | 2.5 | IS-SM 30:1 | ESI (+) | 90 | 10 | 40 | 15 |
| 792.7 | 493.4 | 12 | IS-TG 46:2 | ESI (+) | 130 | 10 | 35 | 10 |
| 496.4 | 184 | 0.9 | LPC 16:0 | ESI (+) | 90 | 10 | 40 | 15 |
| 494.4 | 184 | 0.8 | LPC 16:1 | ESI (+) | 90 | 10 | 40 | 15 |
| 508.5 | 184 | 0.8 | LPC 17:1 | ESI (+) | 90 | 10 | 40 | 15 |
| 524.5 | 184 | 1.2 | LPC 18:0 | ESI (+) | 90 | 10 | 40 | 15 |
| 522.5 | 184 | 0.9 | LPC 18:1 | ESI (+) | 90 | 10 | 40 | 15 |
| 520.5 | 184 | 0.8 | LPC 18:2 | ESI (+) | 90 | 10 | 40 | 15 |
| 518.6 | 184 | 0.7 | LPC 18:3 | ESI (+) | 90 | 10 | 40 | 15 |

| | | | | | | | | |
|---|---|---|---|---|---|---|---|---|
| 546.4 | 184 | 0.9 | LPC 20:3 | ESI (+) | 90 | 10 | 40 | 15 |
| 544.4 | 184 | 0.7 | LPC 20:4 | ESI (+) | 90 | 10 | 40 | 15 |
| 572.4 | 184 | 0.8 | LPC 22:4 | ESI (+) | 90 | 10 | 40 | 15 |
| 570.4 | 184 | 0.8 | LPC 22:5 | ESI (+) | 90 | 10 | 40 | 15 |
| 568.5 | 184 | 0.7 | LPC 22:6 | ESI (+) | 90 | 10 | 40 | 15 |
| 732.6 | 184 | 4.9 | PC 32:1 | ESI (+) | 90 | 10 | 40 | 15 |
| 730.6 | 184 | 4.2 | PC 32:2 | ESI (+) | 90 | 10 | 40 | 15 |
| 728.6 | 184 | 3.6 | PC 32:3 | ESI (+) | 90 | 10 | 40 | 15 |
| 760.6 | 184 | 5.8 | PC 34:1 | ESI (+) | 90 | 10 | 40 | 15 |
| 758.6 | 184 | 5.1 | PC 34:2 | ESI (+) | 90 | 10 | 40 | 15 |
| 756.6 | 184 | 4.6 | PC 34:3 | ESI (+) | 90 | 10 | 40 | 15 |
| 754.6 | 184 | 3.9 | PC 34:4 | ESI (+) | 90 | 10 | 40 | 15 |
| 752.6 | 184 | 3.2 | PC 34:5 | ESI (+) | 90 | 10 | 40 | 15 |
| 788.6 | 184 | 6.8 | PC 36:1 | ESI (+) | 90 | 10 | 40 | 15 |
| 786.6 | 184 | 5.9 | PC 36:2 | ESI (+) | 90 | 10 | 40 | 15 |
| 784.8 | 184 | 5.3 | PC 36:3 | ESI (+) | 90 | 10 | 40 | 15 |
| 782.7 | 184 | 4.9 | PC 36:4 | ESI (+) | 90 | 10 | 40 | 15 |
| 780.8 | 184 | 4.2 | PC 36:5 | ESI (+) | 90 | 10 | 40 | 15 |
| 778.6 | 184 | 3.5 | PC 36:6 | ESI (+) | 90 | 10 | 40 | 15 |
| 776.8 | 184 | 3.6 | PC 36:7 | ESI (+) | 90 | 10 | 40 | 15 |
| 816.6 | 184 | 8.5 | PC 38:1 | ESI (+) | 90 | 10 | 40 | 15 |
| 814.6 | 184 | 7.6 | PC 38:2 | ESI (+) | 90 | 10 | 40 | 15 |
| 812.8 | 184 | 6.6 | PC 38:3 | ESI (+) | 90 | 10 | 40 | 15 |
| 810.8 | 184 | 5.9 | PC 38:4 | ESI (+) | 90 | 10 | 40 | 15 |
| 808.8 | 184 | 5 | PC 38:5 | ESI (+) | 90 | 10 | 40 | 15 |
| 806.7 | 184 | 4.6 | PC 38:6 | ESI (+) | 90 | 10 | 40 | 15 |
| 804.7 | 184 | 3.9 | PC 38:7 | ESI (+) | 90 | 10 | 40 | 15 |
| 844.6 | 184 | 8.8 | PC 40:1 | ESI (+) | 90 | 10 | 40 | 15 |
| 842.6 | 184 | 8.5 | PC 40:2 | ESI (+) | 90 | 10 | 40 | 15 |
| 840.6 | 184 | 7.7 | PC 40:3 | ESI (+) | 90 | 10 | 40 | 15 |
| 838.8 | 184 | 6.7 | PC 40:4 | ESI (+) | 90 | 10 | 40 | 15 |
| 836.8 | 184 | 6 | PC 40:5 | ESI (+) | 90 | 10 | 40 | 15 |
| 834.8 | 184 | 5.6 | PC 40:6 | ESI (+) | 90 | 10 | 40 | 15 |
| 832.7 | 184 | 4.7 | PC 40:7 | ESI (+) | 90 | 10 | 40 | 15 |
| 718.6 | 577.6 | 6.1 | PE 34:1 | ESI (+) | 110 | 10 | 35 | 15 |
| 716.6 | 575.6 | 5.4 | PE 34:2 | ESI (+) | 110 | 10 | 35 | 15 |
| 714.6 | 573.6 | 4.6 | PE 34:3 | ESI (+) | 110 | 10 | 35 | 15 |
| 710.6 | 569.6 | 4.2 | PE 34:5 | ESI (+) | 110 | 10 | 35 | 15 |
| 746.6 | 605.6 | 7.1 | PE 36:1 | ESI (+) | 110 | 10 | 35 | 15 |
| 744.6 | 603.6 | 6.4 | PE 36:2 | ESI (+) | 110 | 10 | 35 | 15 |
| 742.6 | 601.6 | 5.7 | PE 36:3 | ESI (+) | 110 | 10 | 35 | 15 |
| 740.6 | 599.6 | 5.2 | PE 36:4 | ESI (+) | 110 | 10 | 35 | 15 |
| 738.5 | 597.5 | 4.5 | PE 36:5 | ESI (+) | 110 | 10 | 35 | 15 |
| 772.6 | 631.6 | 7.3 | PE 38:2 | ESI (+) | 110 | 10 | 35 | 15 |
| 770.6 | 629.6 | 6.9 | PE 38:3 | ESI (+) | 110 | 10 | 35 | 15 |
| 768.6 | 627.6 | 6.2 | PE 38:4 | ESI (+) | 110 | 10 | 35 | 15 |
| 766.6 | 625.6 | 5.5 | PE 38:5 | ESI (+) | 110 | 10 | 35 | 15 |

| | | | | | | | | |
|---|---|---|---|---|---|---|---|---|
| 764.6 | 623.6 | 4.9 | PE 38:6 | ESI (+) | 110 | 10 | 35 | 15 |
| 762.6 | 621.6 | 4.1 | PE 38:7 | ESI (+) | 110 | 10 | 35 | 15 |
| 796.6 | 655.6 | 6.9 | PE 40:4 | ESI (+) | 110 | 10 | 35 | 15 |
| 794.6 | 653.6 | 6.2 | PE 40:5 | ESI (+) | 110 | 10 | 35 | 15 |
| 792.6 | 651.6 | 5.9 | PE 40:6 | ESI (+) | 110 | 10 | 35 | 15 |
| 790.6 | 649.6 | 5 | PE 40:7 | ESI (+) | 110 | 10 | 35 | 15 |
| 788.6 | 647.6 | 4.4 | PE 40:8 | ESI (+) | 110 | 10 | 35 | 15 |
| 703.7 | 184 | 4.5 | SM 34:1 | ESI (+) | 90 | 10 | 40 | 15 |
| 731.8 | 184 | 5.5 | SM 36:1 | ESI (+) | 90 | 10 | 40 | 15 |
| 759.8 | 184 | 6.6 | SM 38:1 | ESI (+) | 90 | 10 | 40 | 15 |
| 787.8 | 184 | 7.6 | SM 40:1 | ESI (+) | 90 | 10 | 40 | 15 |
| 815.8 | 184 | 8.7 | SM 42:1 | ESI (+) | 90 | 10 | 40 | 15 |
| 813.8 | 184 | 7.6 | SM 42:2 | ESI (+) | 90 | 10 | 40 | 15 |
| 705.5 | 184 | 4.9 | dhSM 34:0 | ESI (+) | 106 | 10 | 35 | 15 |
| 733.5 | 184 | 5.9 | dhSM 36:0 | ESI (+) | 106 | 10 | 35 | 15 |
| 761.5 | 184 | 7 | dhSM 38:0 | ESI (+) | 106 | 10 | 35 | 15 |
| 789.5 | 184 | 8 | dhSM 40:0 | ESI (+) | 106 | 10 | 35 | 15 |
| 817.5 | 184 | 8.8 | dhSM 42:0 | ESI (+) | 106 | 10 | 35 | 15 |
| 815.5 | 184 | 8 | dhSM 42:1 | ESI (+) | 106 | 10 | 35 | 15 |
| 822.6 | 523.6 | 13 | TG 48:1 | ESI (+) | 130 | 10 | 35 | 10 |
| 820.6 | 521.6 | 12 | TG 48:2 | ESI (+) | 130 | 10 | 35 | 10 |
| 850.6 | 551.6 | 13 | TG 50:1 | ESI (+) | 130 | 10 | 35 | 10 |
| 848.6 | 549.6 | 13 | TG 50:2 | ESI (+) | 130 | 10 | 35 | 10 |
| 846.8 | 547.8 | 12 | TG 50:3 | ESI (+) | 130 | 10 | 35 | 10 |
| 878.8 | 579.8 | 14 | TG 52:1 | ESI (+) | 130 | 10 | 35 | 10 |
| 876.8 | 577.8 | 13 | TG 52:2 | ESI (+) | 130 | 10 | 35 | 10 |
| 874.6 | 575.6 | 13 | TG 52:3 | ESI (+) | 130 | 10 | 35 | 10 |
| 872.8 | 573.8 | 12 | TG 52:4 | ESI (+) | 130 | 10 | 35 | 10 |
| 906.8 | 607.8 | 14 | TG 54:1 | ESI (+) | 130 | 10 | 35 | 10 |
| 904.8 | 605.8 | 14 | TG 54:2 | ESI (+) | 130 | 10 | 35 | 10 |
| 902.8 | 603.8 | 13 | TG 54:3 | ESI (+) | 130 | 10 | 35 | 10 |
| 900.7 | 601.7 | 13 | TG 54:4 | ESI (+) | 130 | 10 | 35 | 10 |
| 898.8 | 599.8 | 12 | TG 54:5 | ESI (+) | 130 | 10 | 35 | 10 |
| 928.8 | 629.8 | 13 | TG 56:4 | ESI (+) | 130 | 10 | 35 | 10 |
| 926.8 | 627.8 | 13 | TG 56:5 | ESI (+) | 130 | 10 | 35 | 10 |
| 369.1 | 160.9 | 4.1 | FC | APCI (+) | 90 | 10 | 30 | 15 |
| 376.1 | 160.9 | 4.1 | IS-FCd7 | APCI (+) | 90 | 10 | 30 | 15 |
| 650.1 | 376 | 13.2 | IS-CE 16:0 (d7) | ESI (+) | 81 | 10 | 19 | 10 |
| 642.8 | 369 | 13.2 | CE 16:0 | ESI (+) | 81 | 10 | 19 | 10 |
| 640.8 | 369 | 12.7 | CE 16:1 | ESI (+) | 81 | 10 | 19 | 10 |
| 670.9 | 369 | 13.7 | CE 18:0 | ESI (+) | 81 | 10 | 19 | 10 |
| 668.9 | 369 | 13.2 | CE 18:1 | ESI (+) | 81 | 10 | 19 | 10 |
| 666.8 | 369 | 12.7 | CE 18:2 | ESI (+) | 81 | 10 | 19 | 10 |
| 664.8 | 369 | 12.1 | CE 18:3 | ESI (+) | 81 | 10 | 19 | 10 |
| 698.7 | 369 | 14.1 | CE 20:0 | ESI (+) | 81 | 10 | 19 | 10 |

| | | | | | | | | |
|---|---|---|---|---|---|---|---|---|
| 696.8 | 369 | 13.7 | CE 20:1 | ESI (+) | 81 | 10 | 19 | 10 |
| 692.8 | 369 | 12.8 | CE 20:3 | ESI (+) | 81 | 10 | 19 | 10 |
| 690.8 | 369 | 12.4 | CE 20:4 | ESI (+) | 81 | 10 | 19 | 10 |
| 688.8 | 369 | 11.9 | CE 20:5 | ESI (+) | 81 | 10 | 19 | 10 |
| 726.7 | 369 | 14.4 | CE 22:0 | ESI (+) | 81 | 10 | 19 | 10 |
| 718.8 | 369 | 12.9 | CE 22:4 | ESI (+) | 81 | 10 | 19 | 10 |
| 716.8 | 369 | 12.6 | CE 22:5 | ESI (+) | 81 | 10 | 19 | 10 |
| 714.8 | 369 | 12.2 | CE 22:6 | ESI (+) | 81 | 10 | 19 | 10 |

### REFERENCES

1. Tsuchida T, Lee YA, Fujiwara N, Ybanez M, Allen B, Martins S, et al. A simple diet- and chemical-induced murine NASH model with rapid progression of steatohepatitis, fibrosis and liver cancer. Journal of hepatology. 2018;69(2):385-95.
2. Brunt EM, Janney CG, Di Bisceglie AM, Neuschwander-Tetri BA, Bacon BR. Nonalcoholic steatohepatitis: a proposal for grading and staging the histological lesions. Am J Gastroenterol. 1999;94(9):2467-74.
3. Bedossa P, Poitou C, Veyrie N, Bouillot JL, Basdevant A, Paradis V, et al. Histopathological algorithm and scoring system for evaluation of liver lesions in morbidly obese patients. Hepatology (Baltimore, Md). 2012;56(5):1751-9.
4. Pfaffl MW. A new mathematical model for relative quantification in real-time RT-PCR. Nucleic Acids Res. 2001;29(9):e45.
5. Andrikopoulos S, Blair AR, Deluca N, Fam BC, Proietto J. Evaluating the glucose tolerance test in mice. American journal of physiology Endocrinology and metabolism. 2008;295(6):E1323-32.
6. Babiy B, Busto R, Pastor O. A normalized signal calibration with a long-term reference improves the robustness of RPLC-MRM/MS lipidomics in plasma. Analytical and bioanalytical chemistry. 2021;413(15):4077-90.
7. Folch J, Lees M, Sloane Stanley GH. A simple method for the isolation and purification of total lipides from animal tissues. The Journal of biological chemistry. 1957;226(1):497-509.
8. Peng B, Ahrends R. Adaptation of Skyline for Targeted Lipidomics. Journal of proteome research. 2016;15(1):291-301.
9. Peng B, Kopczynski D, Pratt BS, Ejsing CS, Burla B, Hermansson M, et al. LipidCreator workbench to probe the lipidomic landscape. Nature communications. 2020;11(1):2057.
10. Initiative LS. https://lipidomics-standards-initiative.org/guidelines/lipid-species-quantification. 2020.
